# Polygenic risk vectors (PRV) improve genetic risk stratification for cardio-metabolic diseases

**DOI:** 10.1101/2022.03.02.22271425

**Authors:** Ruowang Li, Xinyuan Zhang, Binglan Li, Qiping Feng, Leah Kottyan, Yuan Luo, Konrad Teodor Sawicki, Atlas Khan, Nita Limdi, Megan Puckelwartz, Wei-Qi Wei, Chunhua Weng, Yong Chen, Marylyn D. Ritchie, Jason H. Moore

## Abstract

Accurate disease risk stratification can lead to more precise and personalized prevention and treatment of diseases. As an important component to disease risk, genetic risk factors can be utilized as an early and stable predictor for disease onset. Recently, the polygenic risk score (PRS) method has combined the effects from hundreds to millions of single nucleotide polymorphisms (SNPs) into a score that can be used for genetic risk stratification. However, current PRS approaches only utilize the additive associations between SNPs and disease risk in a one-dimensional score. Here, we show that leveraging multiple types of genetic effects in multi-dimensional risk vectors, or a polygenic risk vector (PRV), can improve the stratification of cardio-metabolic diseases risks. Using data from UK Biobank (UKBB) and Electronic Medical Records and Genomics (eMERGE) Network biobank linked electronic health records (EHR) as development and evaluation data, we found that the combined effects between the additive PRS and the dominant PRS outperformed either one in terms of disease risk stratification, especially for the individuals in the high-risk group. Our results demonstrate that disease risks are likely to be influenced by multiple types of genetic effects, and PRV could utilize these effects for better risk stratification while retaining the simplicity of the PRS method.

## 2. Introduction

Accurate genetic risk prediction and stratification can play important roles in advanced disease prevention and treatment strategies for hereditary diseases. For most complex diseases, multiple genetic loci encoding numerous genes have been shown to be associated with disease risks^1,2^. However, most individual genetic loci identified in population cohort studies, such as genome-wide association studies (GWAS), have displayed only modest ability to infer a person’s disease risk^3,4^. Recently, the polygenic risk score (PRS) method has improved disease risk stratification performance by aggregating genotype and association information from many to all genetic loci across the genome^5,6^. As a result, numerous methods have been developed and applied to improve the performance of PRS^7–11^.

While empirical evidence has demonstrated the PRS’s improved prediction performance compared to individual genetic loci, few have investigated whether the PRS is the optimal representation of genetic information. In short, a PRS is a linear weighted average of effects from many genetic loci into one variable, which can then be used for disease risk stratification^12,13^. The simplicity of the PRS allows it to be easily constructed and interpreted; however, it omits the potential complexity of the underlying biology. Thus, there is a need for tools that can maintain the simplicity and interpretability of the PRS, at the same time, better incorporate different types of genetic associations.

A common practice for the current PRS is that genetic variations relate to phenotype outcomes through the additive model. This assumption is due to that most existing GWAS assume the additive model as it has higher power to detect statistical associations than other genetic models ^14–16^. Thus, the association coefficients and the SNPs’ allele dosage used to construct PRS are both generated under the additive model. However, as the primary utility of a PRS is towards risk stratification rather than signal detection, the additive PRS alone may not be the most optimal representation of the genetic information. First, the additive model has been commonly used when the true model has not been formally established. Notably, associations identified with an additive model can be better represented by other types of genetic effects^17^. Second, prediction models can include all relevant information to increase the prediction performance as they are not influenced by factors such as multiple testing adjustments^18^. As a result, we propose polygenic risk vector (PRV) that aims to improve disease risk stratification by incorporating information from multiple types of genetic associations while maintaining the simplicity and interpretability of the existing PRS.

To demonstrate the utility of PRV, we utilized multiple biobank linked EHRs to develop and evaluate this new method^19^. We employed cross-validations in the UK BioBank to develop and optimize PRV for two cardio-metabolic diseases: type 2 diabetes and hypertension. The resulting PRVs were applied to the Electronic Medical Records and Genomics (eMERGE) Network EHR data to assess their ability to stratify subjects into different risk groups. In the eMERGE data, the PRVs showed significantly better performance in disease risk stratification than the additive PRS, particularly for the patients in the high-risk group. Our results demonstrated that utilizing multiple types of genetic signals can improve the stratification of cardio-metabolic disease risks. Furthermore, the PRV approach can be straightforwardly adapted to other polygenic diseases.

## 3. Methods

### 3.1. UK Biobank data

The UK Biobank (http://www.ukbiobank.ac.uk/) publicly released ∼500,000 individuals’ genetic-linked phenotype data (application # 32133). The full characteristics of the dataset have been described previously^20^. Individuals were genotyped on two types of genotype arrays (UK BiLEVE Axiom Array or UK Biobank Axiom Array) across 106 batches and imputed using the merged UK10K and 1000 Genomes phase 3 reference panel. Two levels of quality control were performed on the data. At the SNP level, genotype data were filtered so that only SNPs that have less than 5% missing rate, Hardy-Weinberg equilibrium (HWE) p-value > 1e-10, imputation INFO score > 0.8, minor allele frequency > 1%, and a maximum number of alleles of 2 (i.e. bi-allelic SNPs) were kept for further analysis. At the sample level, several steps of filtering were performed. First, samples that failed UKBiLEVE genotype quality control were removed. Second, individuals with kinship coefficient > 0.088388 with others were deemed to be related. One person within each pair of related individuals was randomly removed. Third, individuals who had mismatched genetic-inferred and self-reported sex were removed. Finally, to ensure maximum sample size of discovery and validation data, only individuals of self-reported White British ethnicity were retained.

### 3.2. eMERGE EHR data

Patients’ clinical and genetic data were obtained from the eMERGE Phase III data^21^. The eMERGE data consists of patients from 11 EHR sites in the US. SNPs were imputed using the Haplotype Reference Consortium 1.1 reference under genome build 37. SNPs were filtered to have MAF > 5%, missing rate < 5%, HWE p-value > 1e-10. Common SNPs between eMERGE and UKBB that passed the filtering step were extracted to perform validation. Patients’ genetic ancestry was determined by jointly estimating their top 20 principal components with the 1000 Genomes data. Then, k-means clustering was used to label eMERGE patients’ genetic ancestry based on the population cluster of the 1000 Genomes cohort that the patient coincides. Samples of European ancestry were extracted to ensure consistency with UKBB data. In total, 70,808 adult individuals of European ancestry from eMERGE phase III were used for the validation analysis.

### 3.3. Phenotype extraction

In UKBB, patients’ T2D case status was determined using patients’ responses with a trained nurse or a count of ICD-10 code of E11.X. Patients with co-occurring type 1 diabetes were excluded from the analysis. Hypertension case was defined as having at least one occurrence of ICD-10 code I10. In the eMERGE data, to be counted as T2D cases requires two or more diagnoses of ICD-9 250.00. Similarly, hypertension requires two or more counts of ICD-9 401.9. Patients with one count of either ICD-9 codes were deemed neither a case nor control and excluded from the analysis.

### 3.4. Development of the polygenic risk vector method

The polygenic risk vector (PRV) is an extension of the polygenic risk score. A PRS is calculated as the sum of SNPs’ association coefficient weighted by the genotype dosage. For N selected SNPs, a PRS is calculated as:

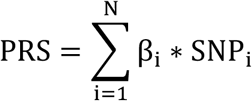

where β_i_ is the effect size of an individual SNP_i_ on a certain phenotype and SNP_i_ is the allelic dosage counts. Commonly, both β_i_ and SNP_i_ in a PRS are assumed to be under the additive model, which implicates a linear relationship between the minor allele counts and the disease risk. However, there are other types of genetic models (Table 1) that can characterize non-additive relationships between a SNP and a phenotype^22,23^.

**Table 1.** SNP coding under different genetic models

| SNP alleles (minor allele = a) | Additive model | Dominant model | Recessive model |
| --- | --- | --- | --- |
| AA | 0 | 0 | 0 |
| Aa | 1 | 1 | 0 |
| aa | 2 | 1 | 1 |

As a result, a PRS can be constructed for every type of genetic model.

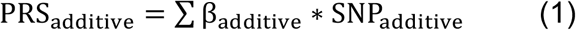

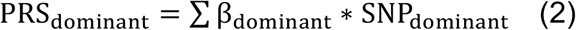

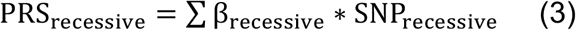

Importantly, PRS constructed under different types of genetic models could provide orthogonal or independent predictions of the underlying genetic risks. Thus, the proposed PRV aims to utilize multiple types of PRS to improve the stratification of disease risks.

We first used the UKBB data to construct and tune the PRV. First, the data was split into 70% training and 30% testing sets. Second, plink 2.0 was used in the training data to generate the genetic association for each SNP under the additive, dominant, and recessive models in each data split. Specifically, SNPs’ associations with the phenotype were determined under the additive, dominant, or recessive model while adjusting for gender and top ten genetic principal components, respectively. Then, the pruning and thresholding method (plink --clump-r2 0.25) with three different p-value thresholds, 0.05, 0.5, and 1, were used to select the SNPs included in the PRS calculation^24^. Thus, a total of 3 (p-value thresholds) x 3 (genetic models) x 2 (diseases) = 18 sets of SNPs with their association coefficients were obtained from the training data. Third, PRS was constructed using SNPs’ coefficients generated from the training data in the testing data. Finally, the entire procedure was repeated for five times, each time using a different data split.

### 3.5. Strategies for tuning PRV

The goal of PRV is to utilize multiple types of PRS to better stratifying patients compared to individual PRS. Thus, increasing the enrichment of diseases cases in the PRV- identified high risk group would be the basis for the tuning of PRV. As the contributions from individual PRSs in a PRV are likely to be non-linear and disease dependent, we propose a data-driven approach to identify differential risk groups in the discovery data and apply the resulting risk groups in the evaluation data.

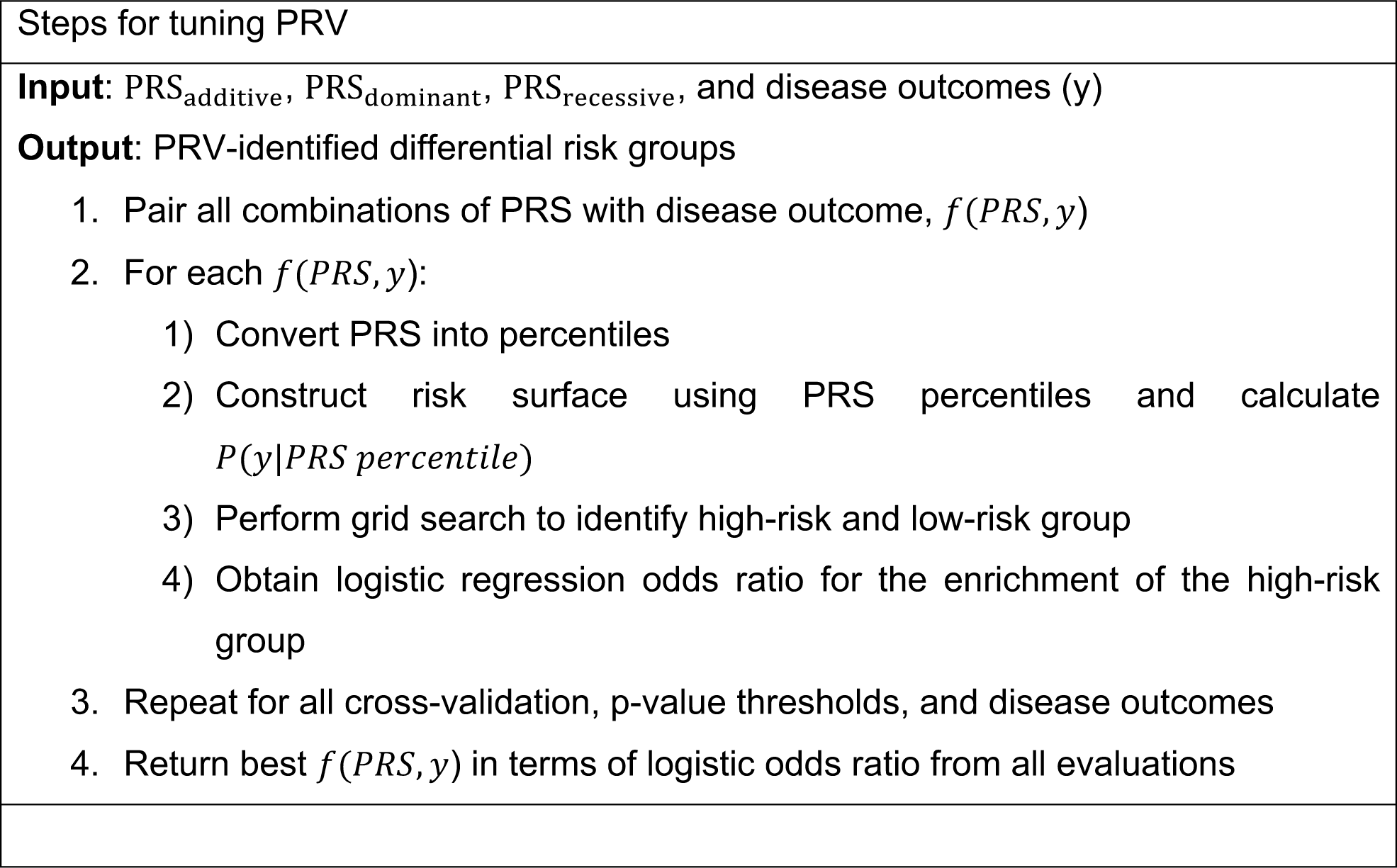

The UKBB testing data was used for the tuning of PRV. For each disease, the best performing PRV in risk stratification was independently evaluated in the eMERGE data.

The capacity of PRV to stratify disease risk was first examined through visualization. For each disease and p-value combination, one-dimensional PRS was first converted into percentiles, and then, at each risk score percentile, the disease prevalence was calculated and averaged across cross-validations. The relationship between the risk score percentile and disease prevalence was visualized. For PRV that consists of two risk scores, both risk scores were first converted into percentiles, and the disease prevalence for the joint distributions were averaged across CVs. For the PRV that included three risk scores, zero counts or low patients count were found in most joint percentile combinations. Thus, PRV containing all three scores was not pursued further (see Discussion).

As a comparison, the same procedure was repeated in the eMERGE data. First, PRV was constructed using association coefficients generated in the UKBB training data. Then, the two-dimensional PRV was visualized in the eMERGE data using its disease prevalence.

To select the best performing PRV, *f*(·) was tuned through a grid search in the UKBB testing data. For one-dimensional risk scores, the percentile of the risk score was treated as grids, which were then ranked by the disease prevalence. The risk stratification performance was determined as the odds ratio of case enrichment between the top 10% grids vs. the bottom 10% grids. The risk surface was evenly divided into 16 × 16 grids for two-dimensional risk vectors, which were then ranked. The performance was similarly determined as the enrichment of the top 10% vs. the bottom 10% grids. Grids that contained less than 50 individuals were dropped to avoid overfitting. Out of all optimization combinations, the PRV with the highest odds ratio enrichment was further evaluated in the eMERGE data.

### 3.6. Evaluating PRV using eMERGE

For each disease, the best one-dimensional PRS and two-dimensional PRV obtained from UKBB were evaluated in the eMERGE data. Each PRS and PRV transferred from UKBB to eMERGE data consists of the association coefficients generated under the additive, dominant, and recessive model, and the optimized *f*(·) ,i.e., the relative ranking of the disease risk grids. eMERGE PRV was then calculated using these summary statistics. The performance of PRV was evaluated as the odds ratio of disease enrichment between the top X% grids vs bottom 10% grids, with X varying from 1 to 50.

## 4. Result

### 4.1. Overview of the analysis

PRV aims to improve the performance of disease risk stratification by utilizing multiple types of genetic effects. In order to demonstrate the effectiveness of the PRV, UKBB data was used for developing and tuning the PRV. The UKBB data were divided into training and testing splits. PRV for T2D and hypertension were generated in the training data, which were then tuned using the testing data split. The best PRVs from the UKBB were evaluated in the eMERGE data (Figure 1).

**Figure 1.**
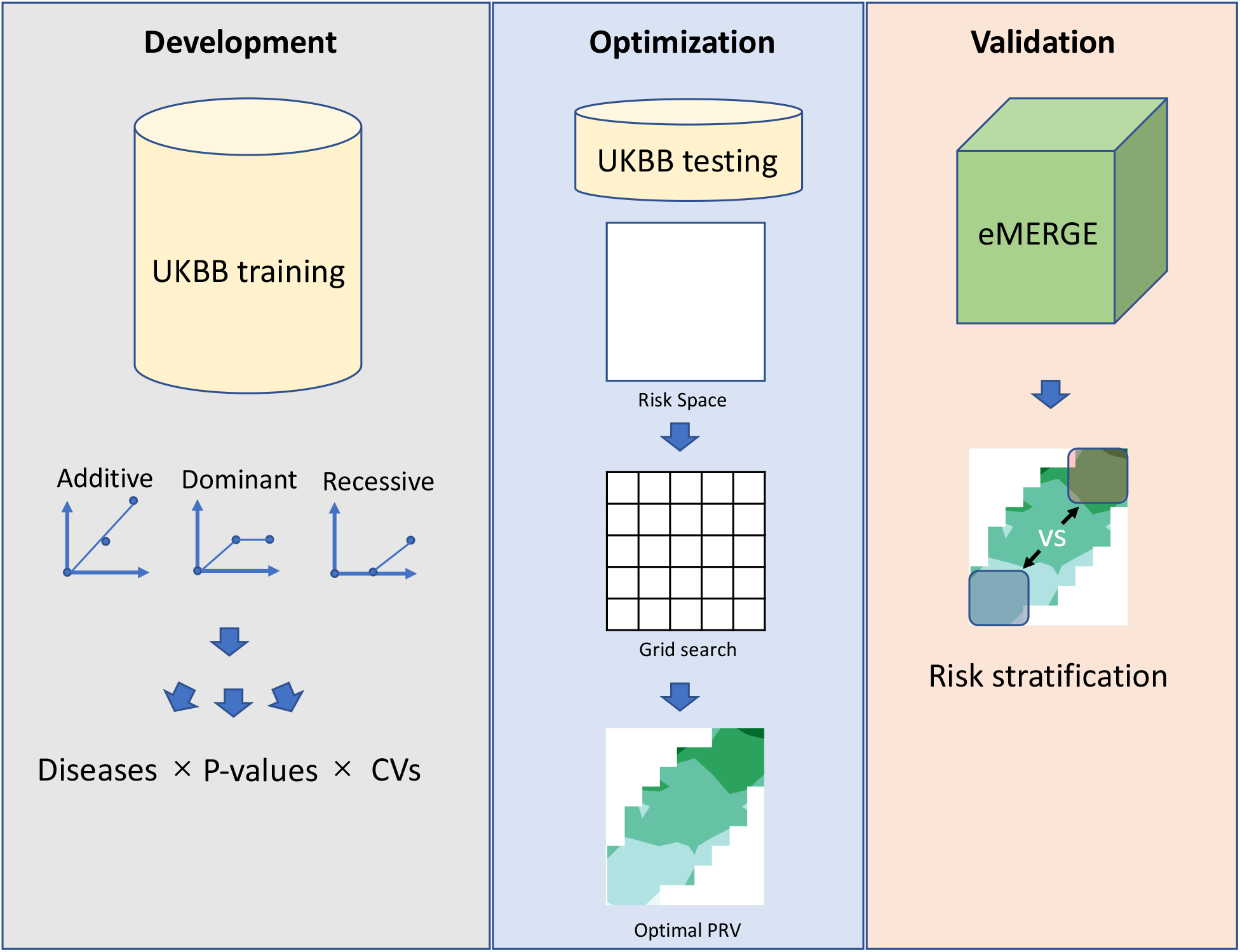
Developing and evaluation scheme for PRV

### 4.2. Individual and joint effects between the additive PRS and dominant PRS

Two cardio-metabolic diseases: T2D and hypertension were extracted from UKBB and eMERGE (Table 2). In the UKBB data, genetic associations for the phenotypes were obtained from the training data, and PRS was constructed in the testing data using the association coefficients.

**Table 2.** Case and control counts for the cardio-metabolic diseases.

|  | Gender | UK BioBank |  | eMERGE Phase III |  |
| --- | --- | --- | --- | --- | --- |
|  |  | Cases | Controls | Cases | Controls |
| Type 2 diabetes | Male | 13,274 | 170,543 | 6,978 | 24,951 |
|  | Female | 8,174 | 206,106 | 5,537 | 30,132 |
| Hypertension | Male | 48,236 | 135,946 | 15,987 | 14,718 |
|  | Female | 40,771 | 173,618 | 15,280 | 19,141 |

For one-dimensional PRS scores, the additive and dominant PRS displayed similar ability for risk stratification in the UKBB testing data. However, the recessive PRS was not correlated with disease risks (Figure 2). Results for other p-value thresholds can be found in the supplemental figures.

**Figure 2.**
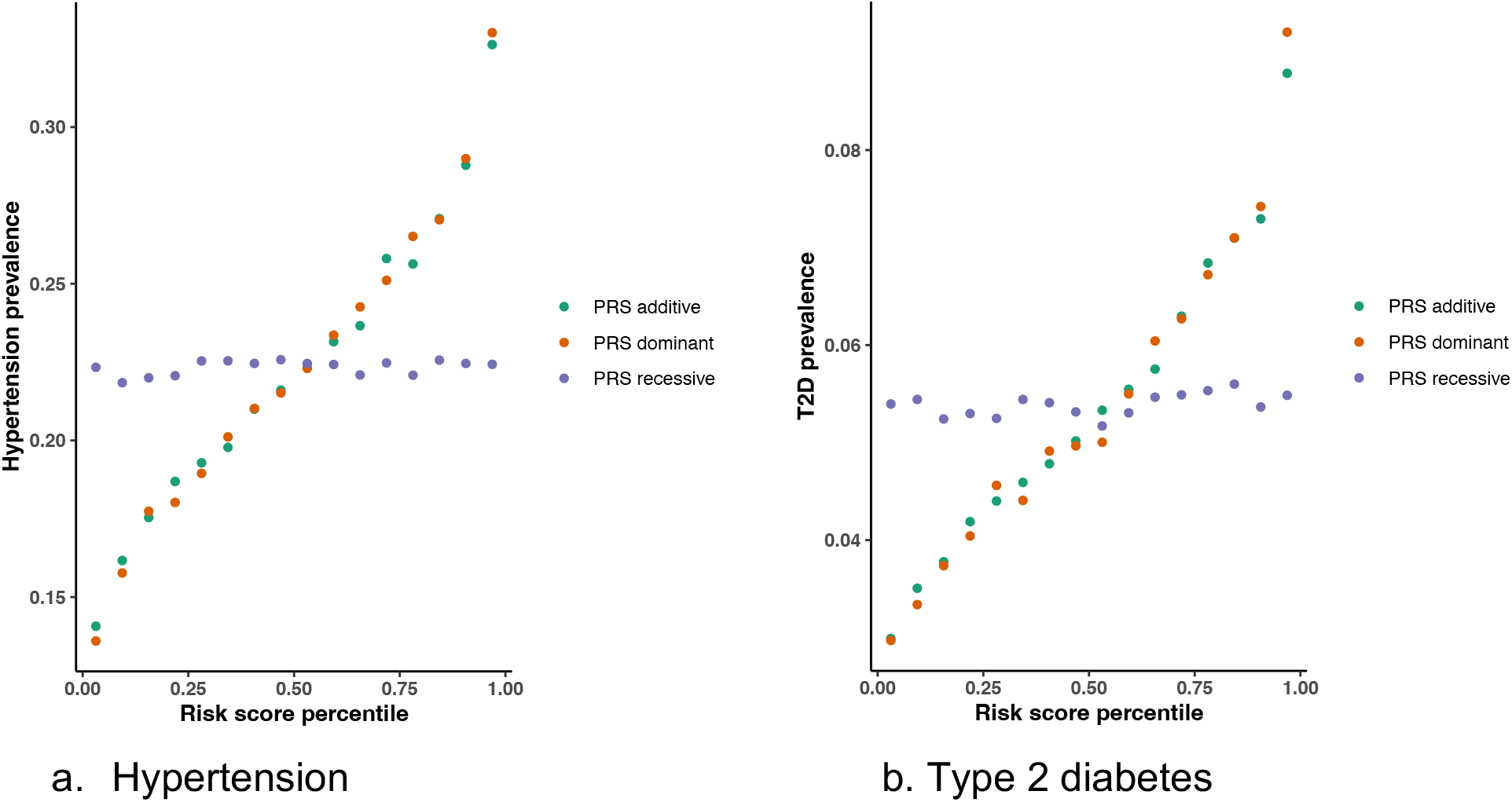
Relationship between PRS risk percentile and disease prevalence. The genetic associations obtained from the UKBB training data were used to calculate the additive, dominant, and recessive PRS in the UKBB testing data. Each disease prevalence was calculated as the number of cases divided by the total individual count at each risk score percentile. SNP p-value threshold = 0.5.

Among the three types of two-dimensional PRVs, the additive PRS and dominant PRS have shown a non-linear modification of disease risk across all SNP selection thresholds. In general, individuals that exhibit high risk in both the additive PRS and dominant PRS have the highest disease prevalence. Conversely, individuals of low risk for both scores have the lowest disease risk. In addition, the recessive PRS did not display noticeable joint effects with other PRS in stratifying the disease risks (Figure 3 and 4). Results for other p-value thresholds can be found in the supplemental figures.

**Figure 3.**
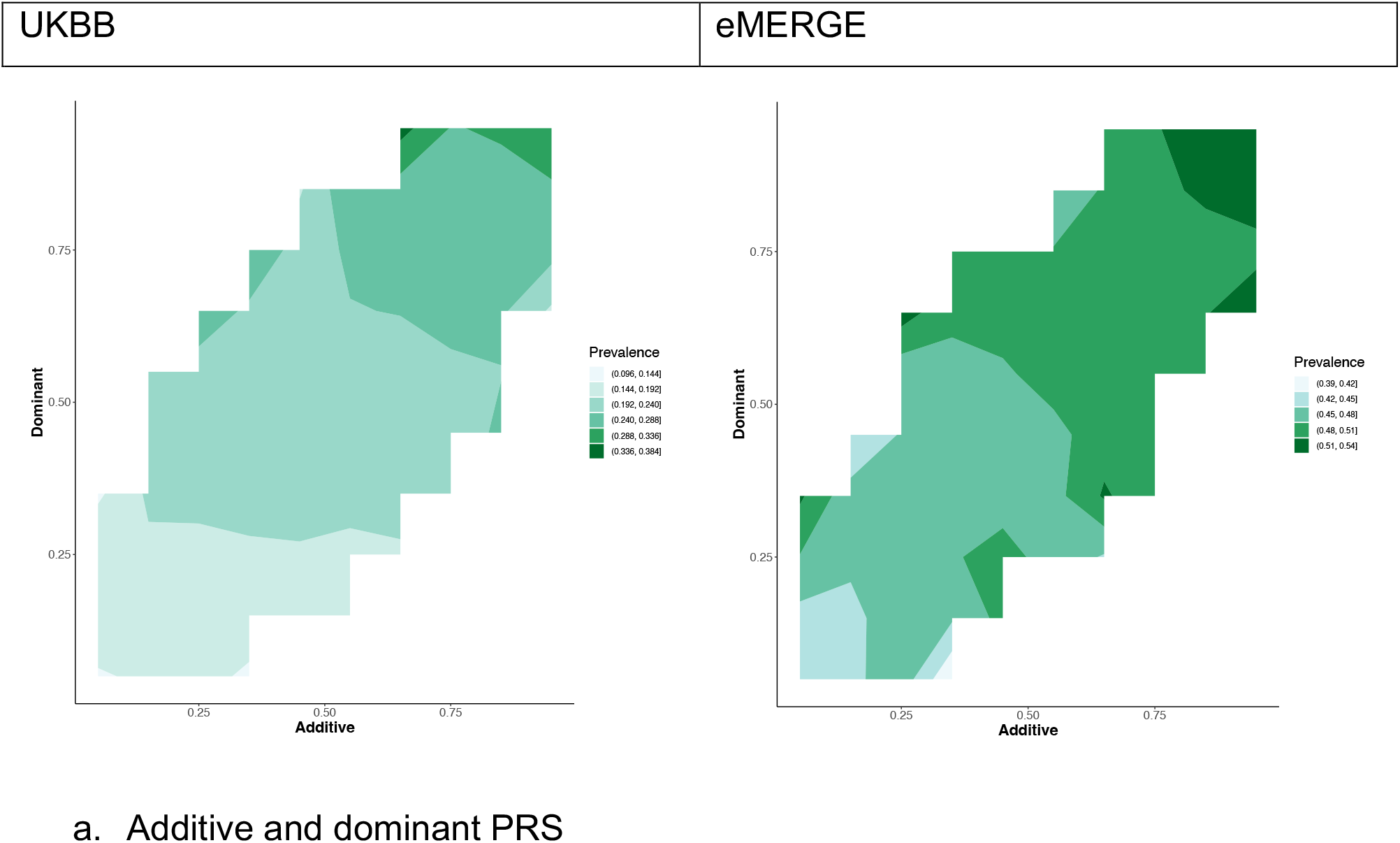
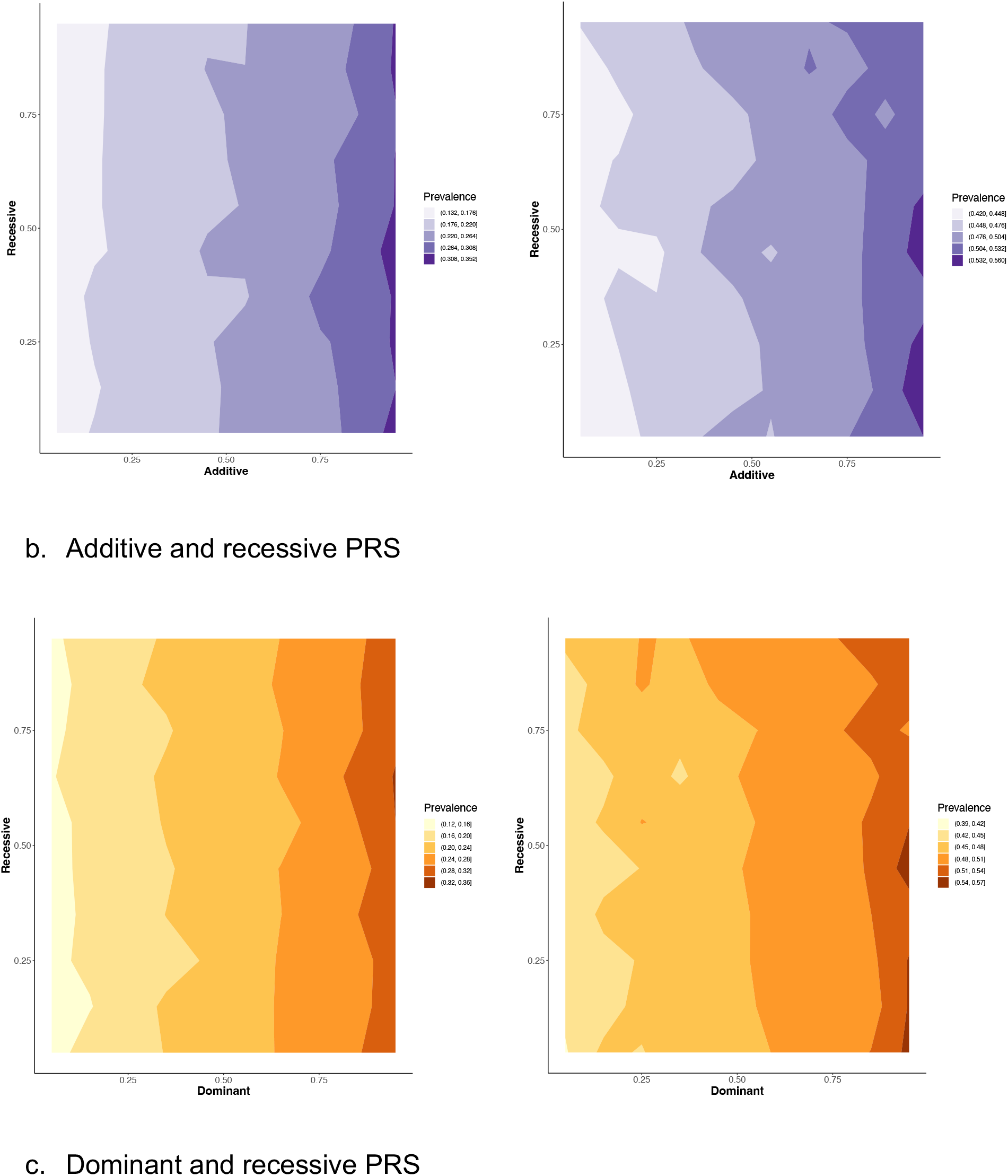
Multi-dimensional PRV and hypertension risk. In each subfigure, two types of PRS risk percentiles were plotted on the x-axis and y-axis, respectively. The disease prevalence at each joint risk percentile was averaged over all cross-validations and displayed as color intensity. Smoothing was applied to discretize disease prevalence for better color separation. Results were based on the SNP p-value threshold of 0.5. Left panel: UKBB. Right panel: eMERGE.

**Figure 4.**
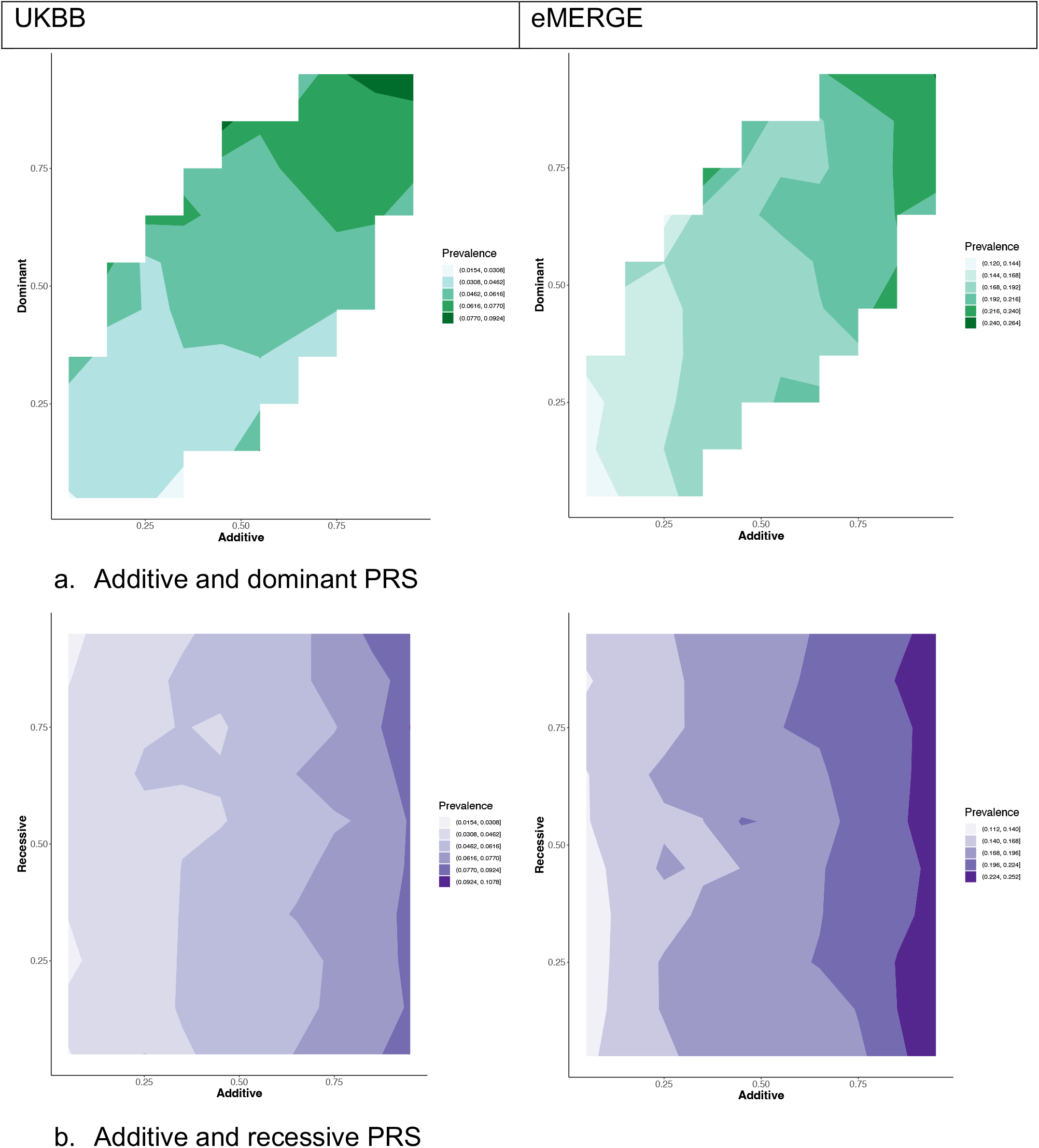
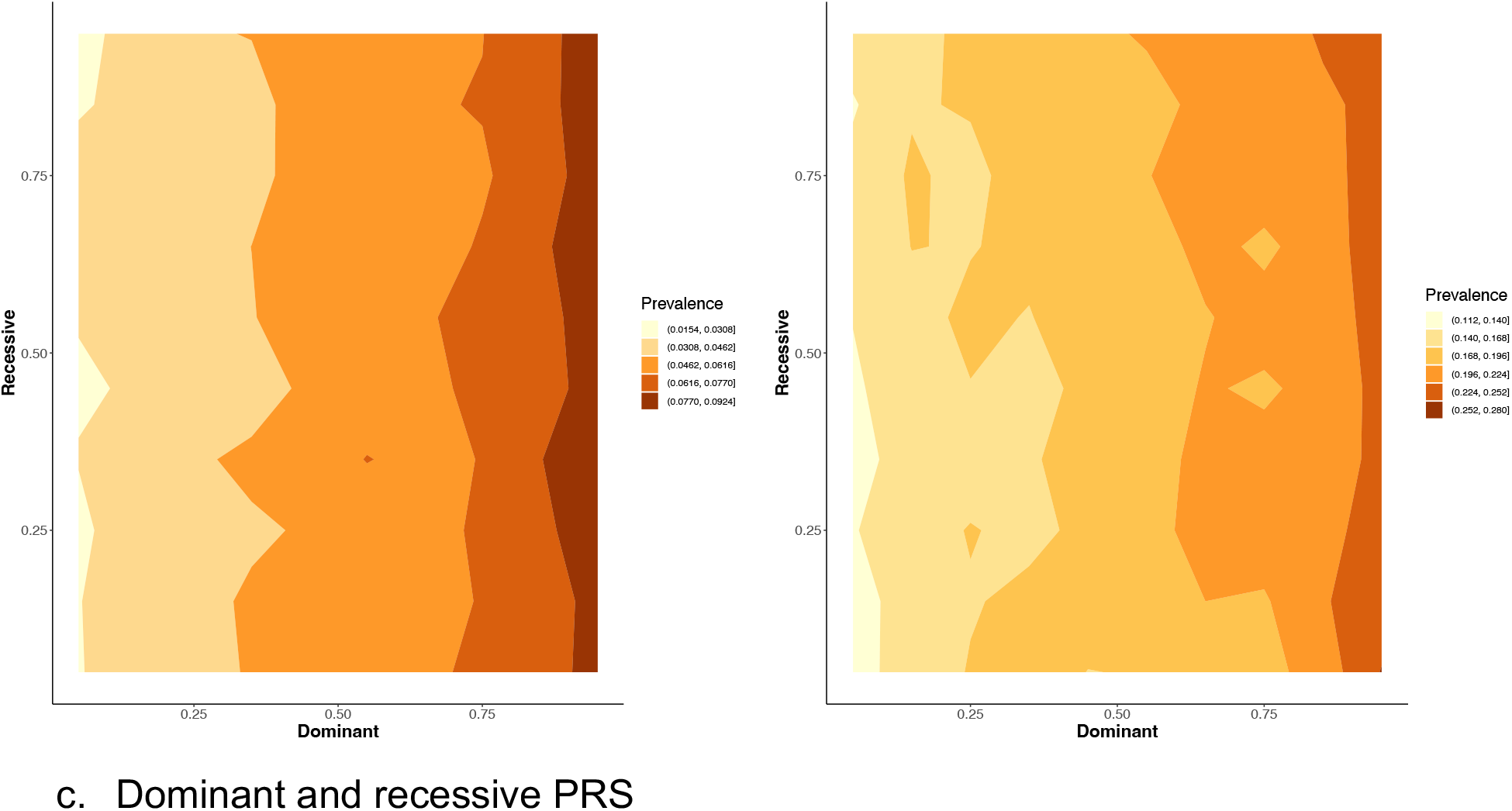
Multi-dimensional PRV and T2D risk. In each subfigure, two types of PRS risk percentiles were plotted on the x-axis and y-axis, respectively. The disease prevalence at each joint risk percentile was averaged over all cross-validations and displayed as color intensity. Smoothing was applied to discretize disease prevalence for better color separation. Results were based on the SNP p-value threshold of 0.5. Left panel: UKBB. Right panel: eMERGE

Grid search was employed to tune PRV in the UKBB testing data. Across all data splits and SNP p-value thresholds, the best performing one-dimensional PRS and two-dimensional PRV were determined for each disease. Because the recessive PRS was not informative in stratifying the disease risk, PRVs that contain recessive PRS were not pursued further.

### 4.3. Risk stratification of T2D and hypertension in eMERGE

The best PRVs from UKBB were evaluated in the eMERGE data to assess their ability for risk stratification. For both T2D and hypertension, PRVs that utilized both additive and dominant genetic effects were better in stratifying disease risks than either additive or dominant PRS alone. The improvement is more pronounced for the highest risk individuals, as the differences were statistically significant (Figure 5).

**Figure 5.**
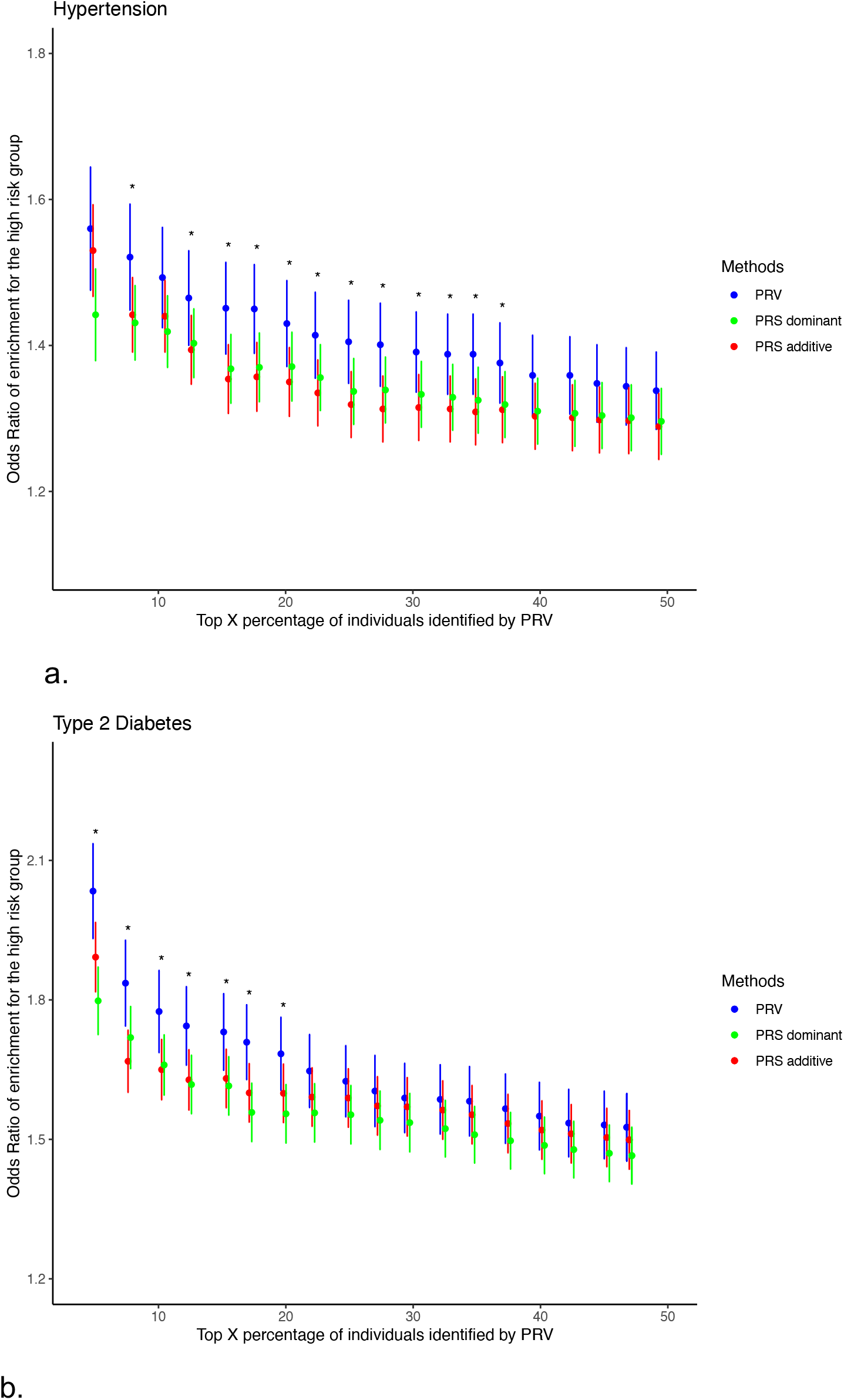
Comparisons of risk stratification between PRV and PRS in eMERGE. The x-axis displays the top X% of individuals, with X varying from 1 to 50, selected by the PRV in terms of disease risk. The specificity decreases from left to right as more lower-risk individuals are included. The y-axis displays the odds ratio enrichment of cases, along with the 95% confidence interval, of the high-risk group. For additive and dominant PRS, the high-risk and the low-risk groups were selected only based on the one-dimensional risk score. Statistical significance was determined using the one-sided Wilcoxon test and denoted by the *. a. Hypertension b. Type 2 diabetes

## 5. Discussion

Genetic risk is a stable and early risk predictor for heritable diseases^25^. As cardio- metabolic diseases are highly heritable, accurate and early risk stratification could lead to better prevention and treatment of the diseases^26^. Thus far, PRS has been widely used as a tool for genetic risk prediction. Numerous methods have been developed to improve the performance of PRS; however, they all fall under the existing PRS framework, which combines SNPs’ additive genetic effects into a one-dimensional score^7–11^. Here, we introduce an orthogonal approach, PRV, to improve genetic risk stratification by utilizing multiple types of genetic effects as multi-dimensional risk vectors.

First, we evaluated the one-dimensional PRS generated under the dominant and recessive models in addition to the additive model. In the UKBB testing data, dominant PRS showed similar performance as the standard additive PRS. However, recessive PRS could not predict the disease risk (Figure 2). The high correlation between the two models could explain the similarity in performance between the additive and dominant PRS (Table 1). On the other hand, recessive effects are known to be associated with many Mendelian diseases^27^. Thus, the recessive PRS could be important for other diseases and thus should be further explored.

For PRVs that utilized two-dimensional genetic effects, the recessive PRS again did not show additional modifying effects on individuals’ genetic risk. This is shown by the vertical or horizontal contours of disease prevalence in Figures 3 and 4. Thus, we did not pursue higher dimensional PRVs involving recessive PRS. In contrast, the additive and dominant PRS jointly influenced the individuals’ genetic risk. The disease prevalence contours were not parallel to the vertical or horizontal axis, which indicates individuals under the same additive genetic risks can be further stratified using the dominant genetic risks, and vice versa. Thus, despite being correlated, the two types of genetic effects have independent predictability on individuals’ cardio-metabolic disease risks.

Through optimization, we selected the best performing PRVs from the UKBB and applied them to the eMERGE data. The result showed that the multi-dimensional PRV outperformed the one-dimensional PRS. Importantly, for the highest risk group, PRV is statistically better at risk stratification than either additive or dominant PRS (Figure 5). The enrichment of the high-risk group has great clinical significance because the specificity of the prediction in a large part determines the feasibility of implementing genetic risk prediction in clinic. As the high-risk groups are more likely to benefit from improved prevention and treatment strategies, improving the identification of this group is crucial. Because PRV incorporates individual PRSs, risk stratification using PRV will consistently perform equal or better than PRS, which indicates that PRV is better suited to identify high-risk individuals than PRS.

Compared to PRS, PRV offers a more generalized framework to utilize genetic information. The additive PRS can be considered a special case of PRV, i.e. one-dimensional additive PRV. However, the common choice of the additive model is due to considerations of power in detecting significant genetic associations. For prediction tasks, power is not of primary concern; thus, all relevant genetic effects should be included. Although an improvement, similar to PRS, PRV is currently limited to provide the relative disease risk of individuals. Further work is needed to integrate PRV with other risk factors to provide the absolute risk of an individual^28–30^. In addition, we have utilized the pruning and thresholding method to select SNPs into the PRV. Under the additive framework, other methods have been developed to better account for linkage-disequilibrium between SNPs. Further work is needed to incorporate this and other features into multi- dimensional risk vectors in PRV. Finally, the PRV can be straightforwardly applied to other polygenic diseases and traits to evaluate the role of recessive genetic effects.

## Data Availability

The UK biobank data can be applied from https://www.ukbiobank.ac.uk/. The eMERGE data can be applied from https://emerge-network.org/

## 6. Acknowledgement

We would like to thank UKBB and eMERGE for providing the data for this study. We would like to acknowledge the grant support from NIH LM010098.

## 7. Author contributions

R.L., J.H.M. devised the project. R.L. performed the analysis and wrote the paper. X.Z., and B.L. assisted the analysis. Y.C. and M.D.R. contributed to the interpretation of results. All authors provided guidance to the project.

