## Supplemental Figures for "Polygenic risk vectors (PRV) improve genetic risk stratification for cardio-metabolic diseases"

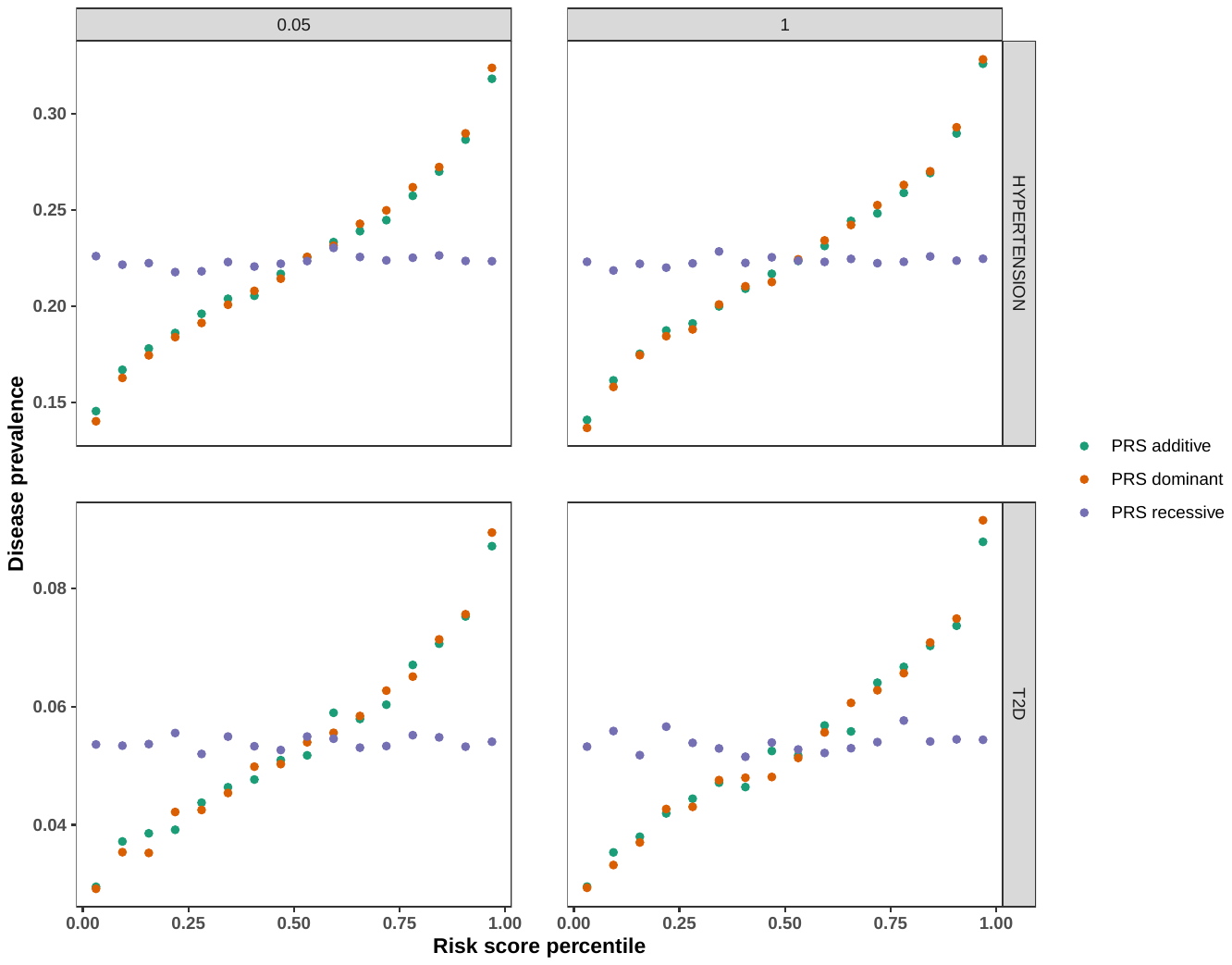

Supplemental Figure 1. Relationship between PRS risk percentile and disease prevalence for additional p-values. In addition to the p-value threshold of 0.5 used in Figure 2. P-value threshold of 0.05 and 1 were used to construct PRS for Hypertension and T2D.

| UKBB | eMERGE |
| --- | --- |
| 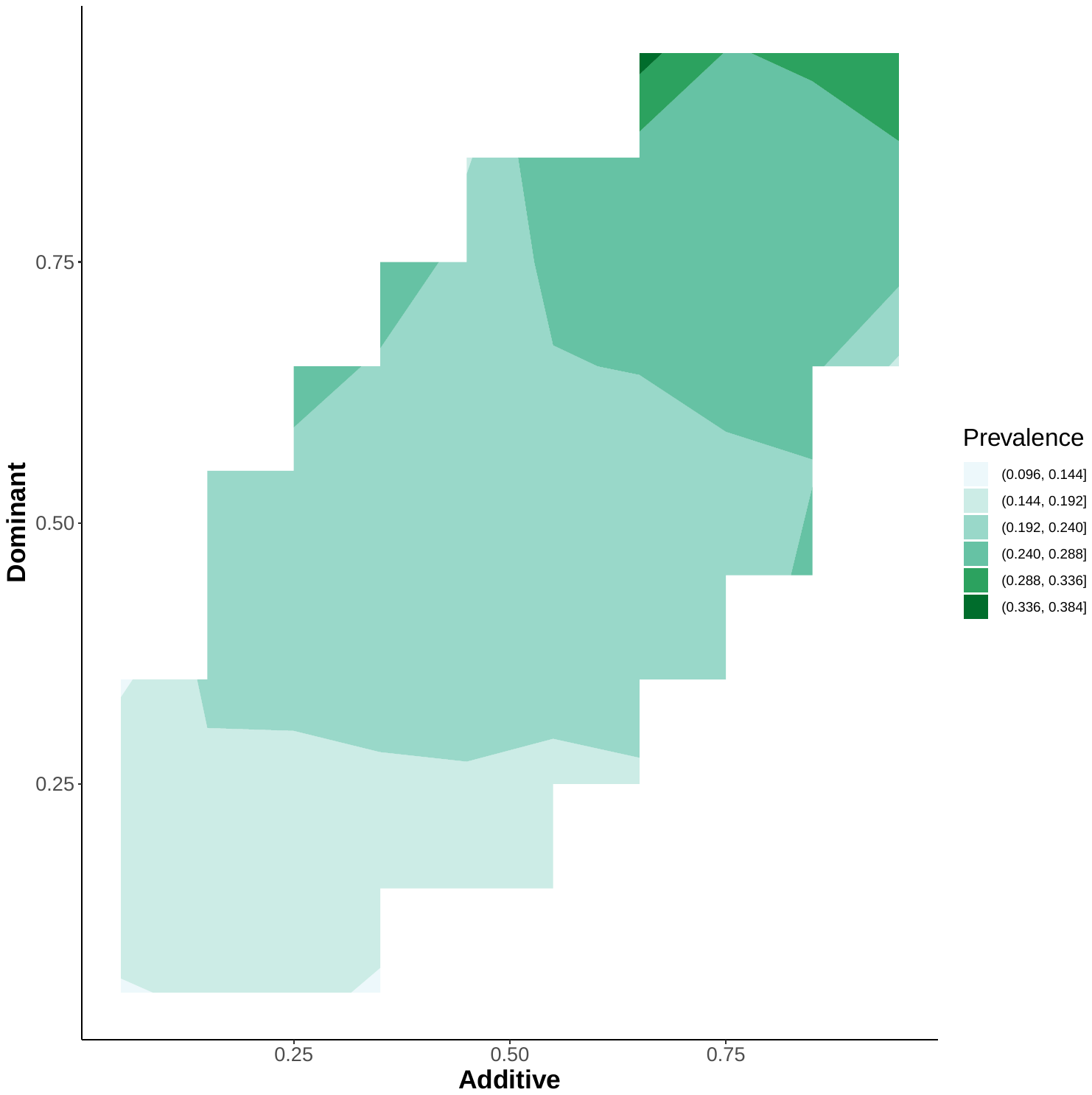 | 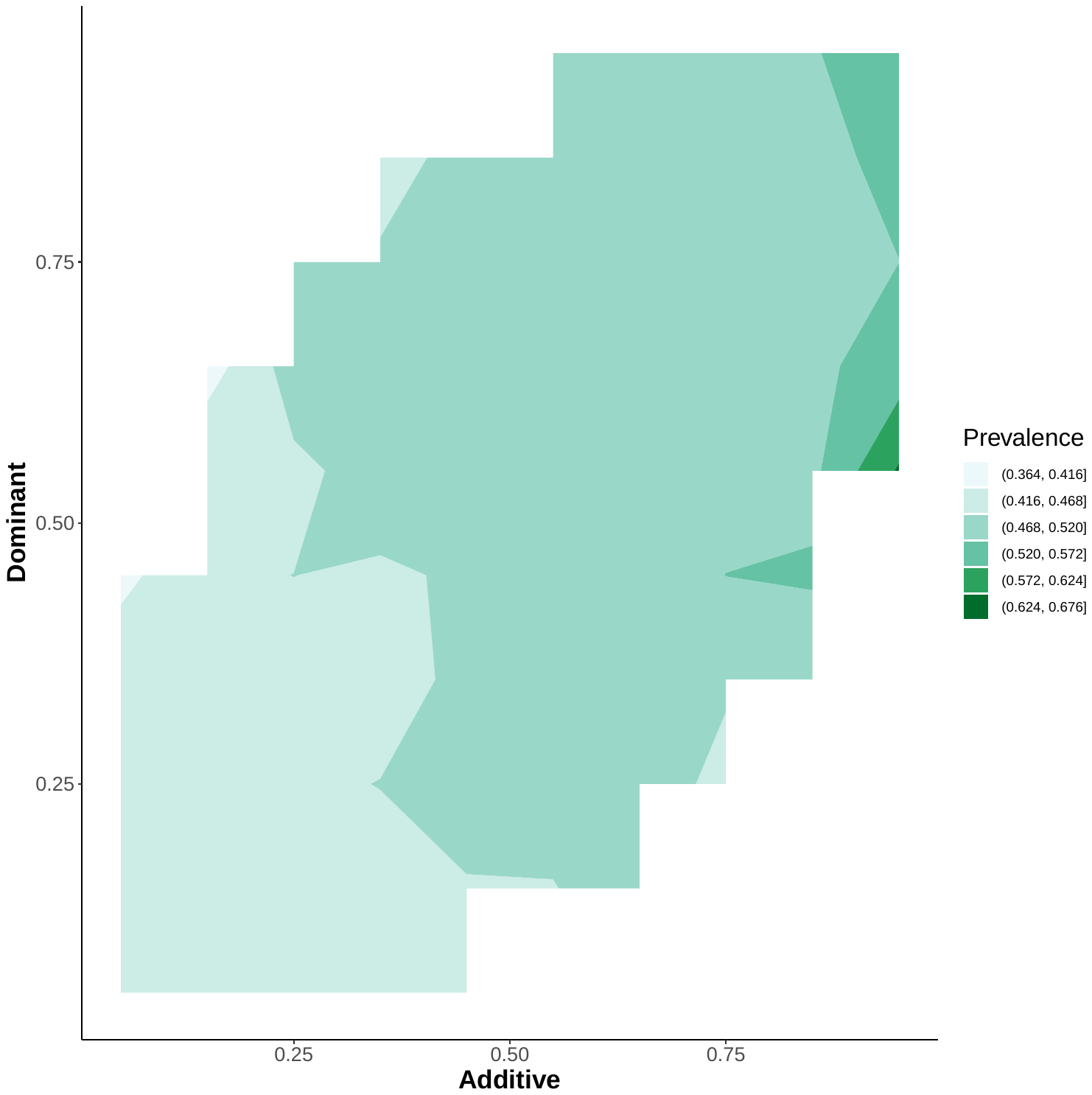 |
| a. Additive and dominant PRS | |
| 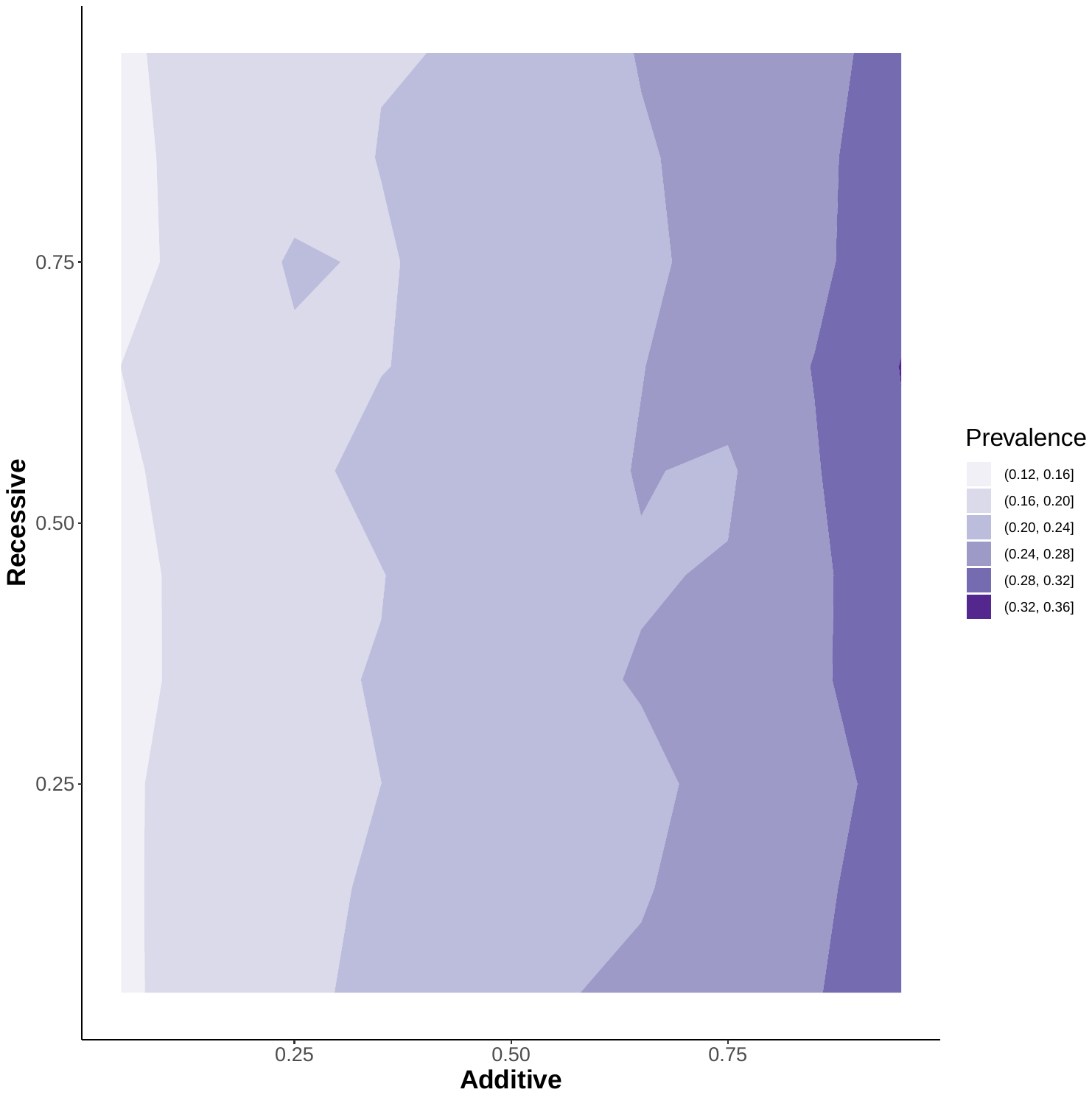 | 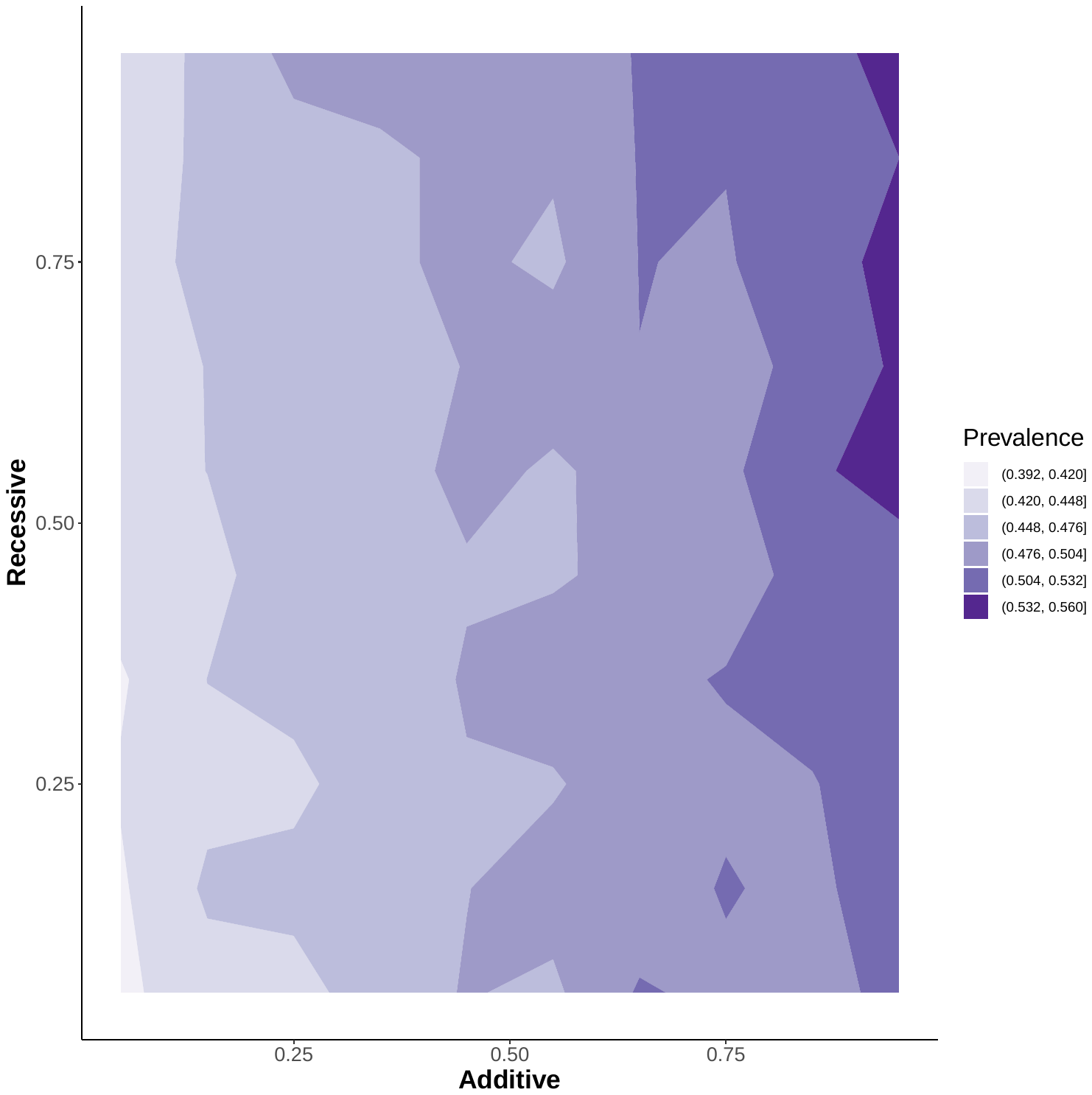 |
| b. Additive and recessive PRS | |
| 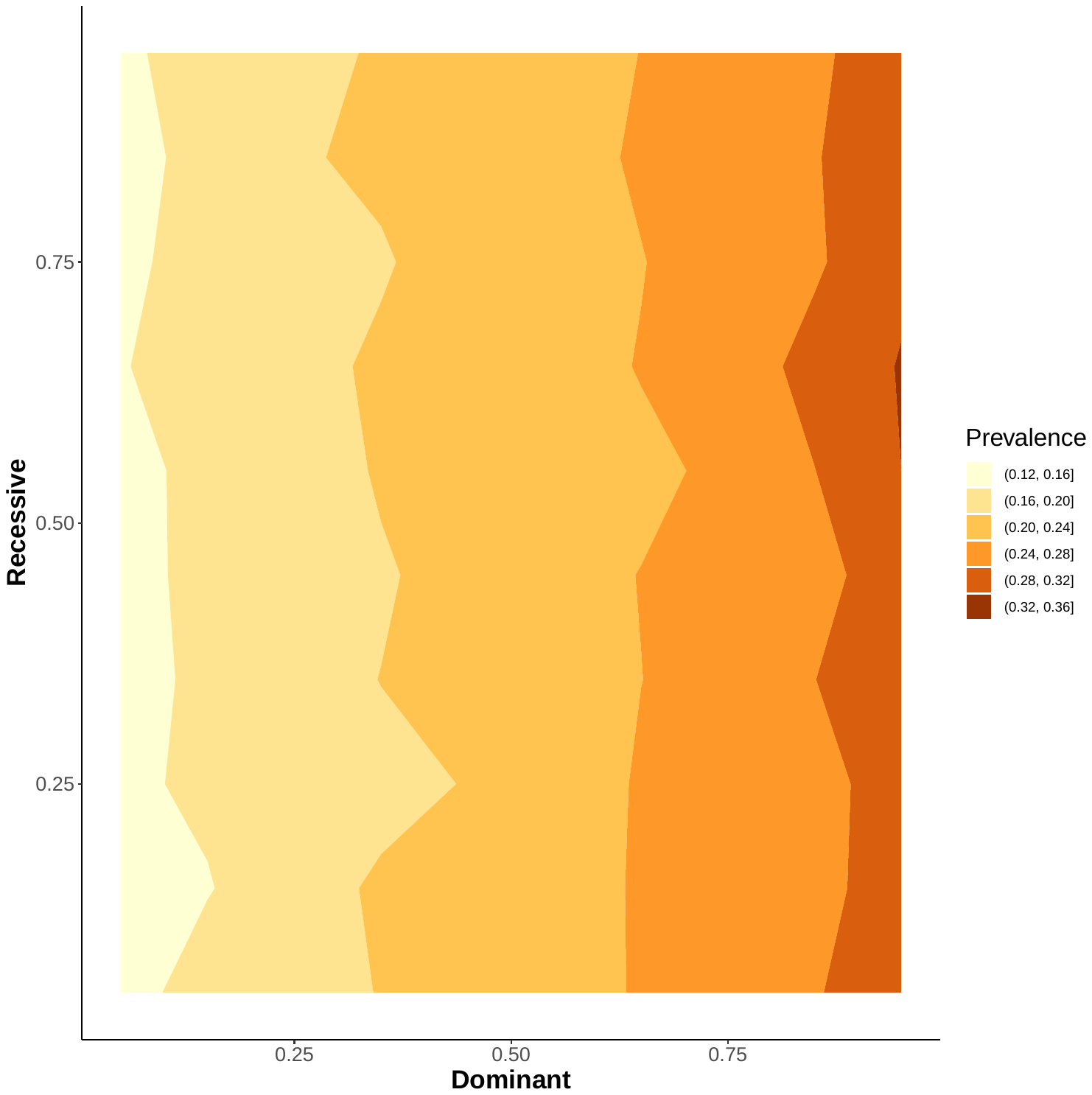 | 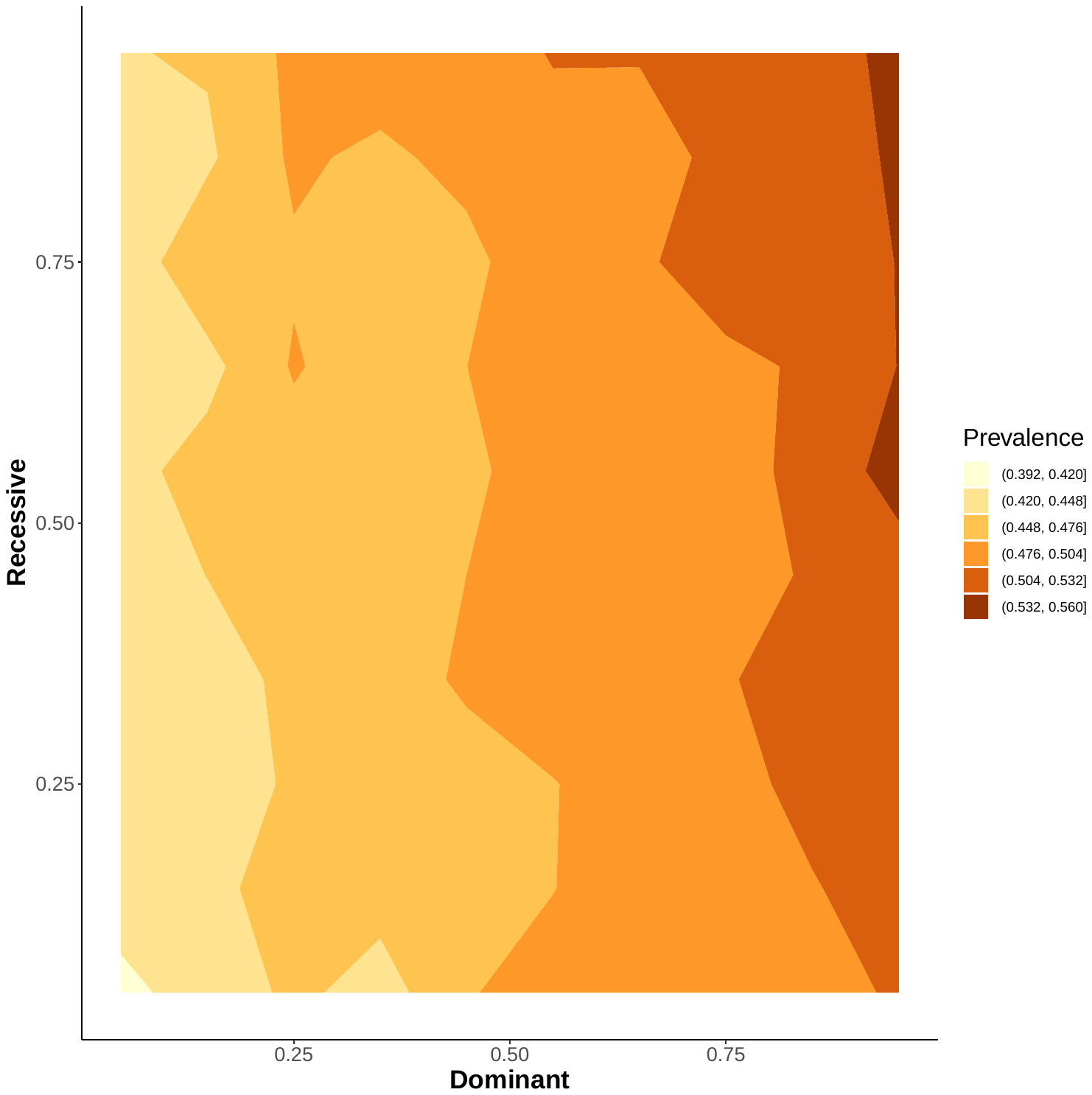 |
| c. Dominant and recessive PRS | |

Supplemental Figure 2. Multi-dimensional PRV and hypertension risk under p-value of 0.05.

| UKBB | eMERGE |
| --- | --- |
| 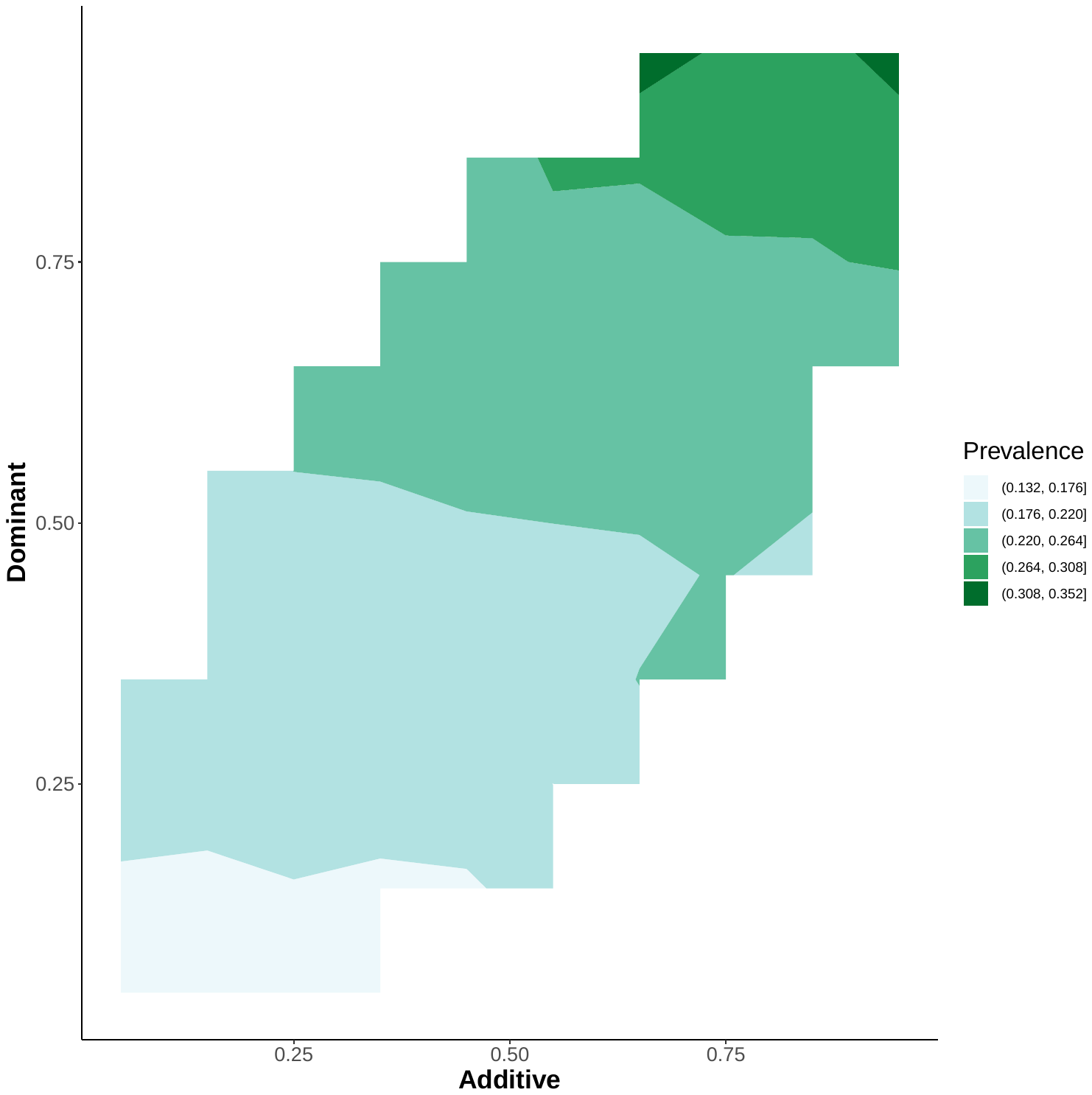 | 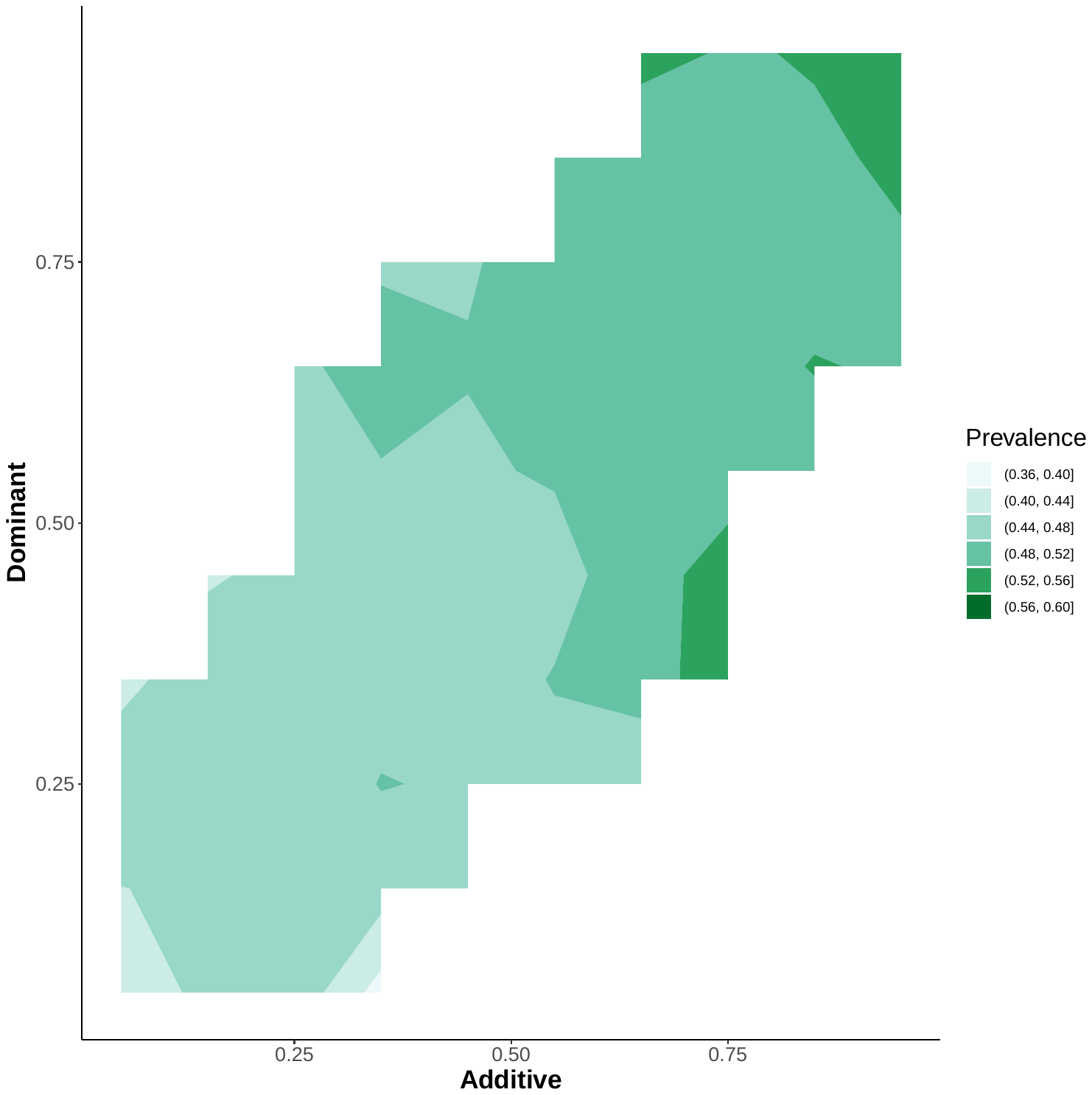 |
| a. Additive and dominant PRS | |
| 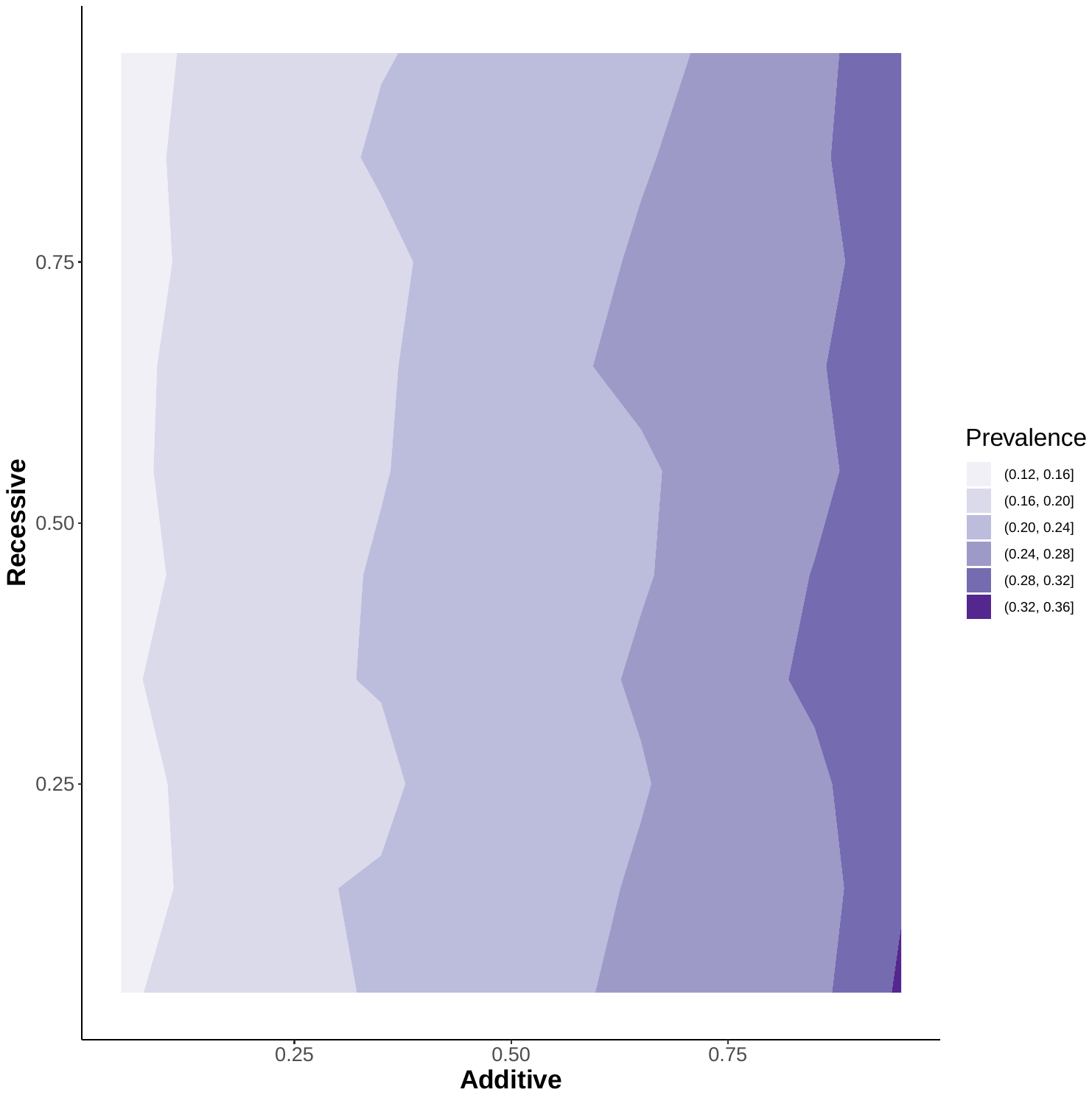 | 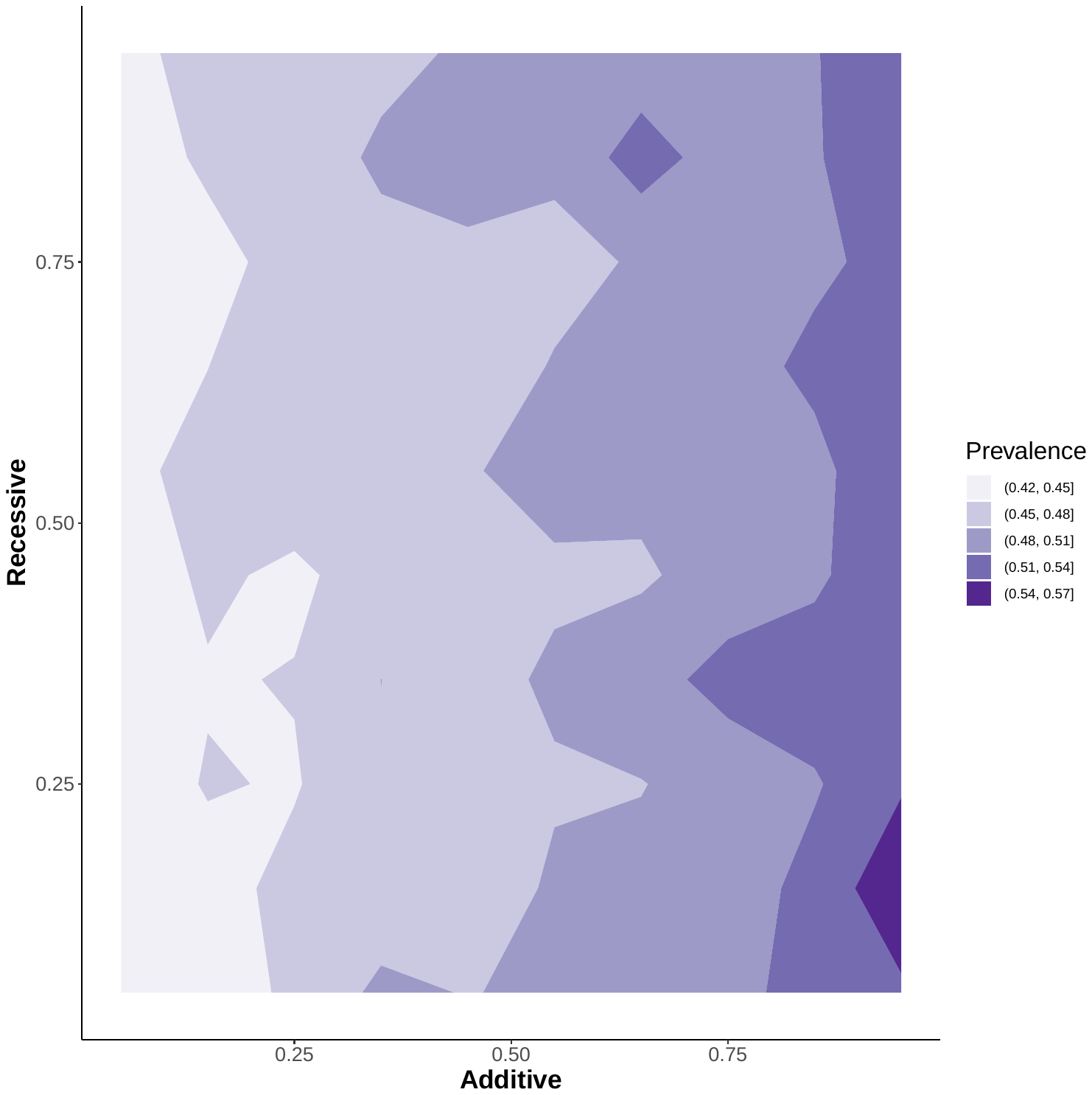 |
| b. Additive and recessive PRS | |
| 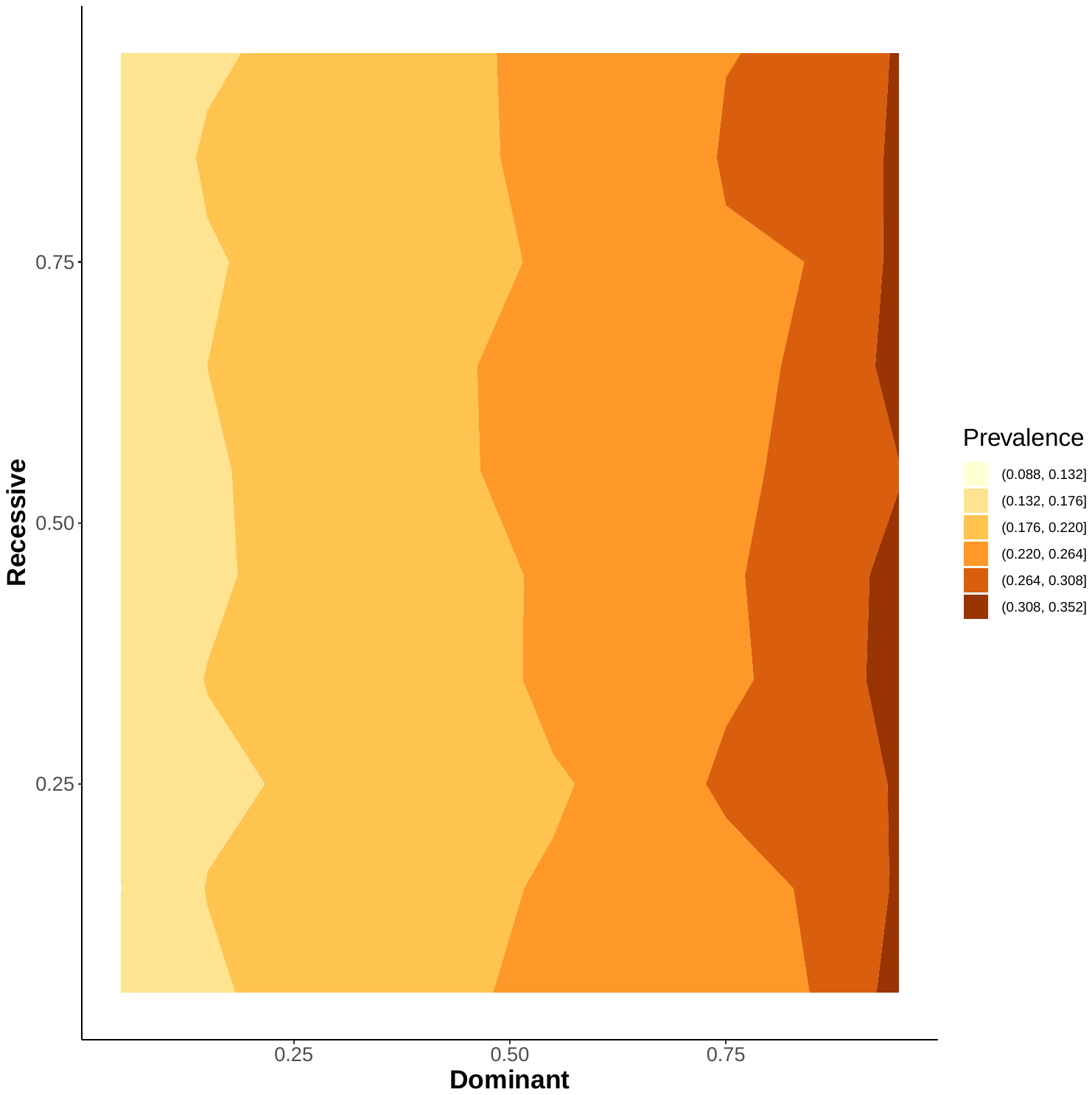 | 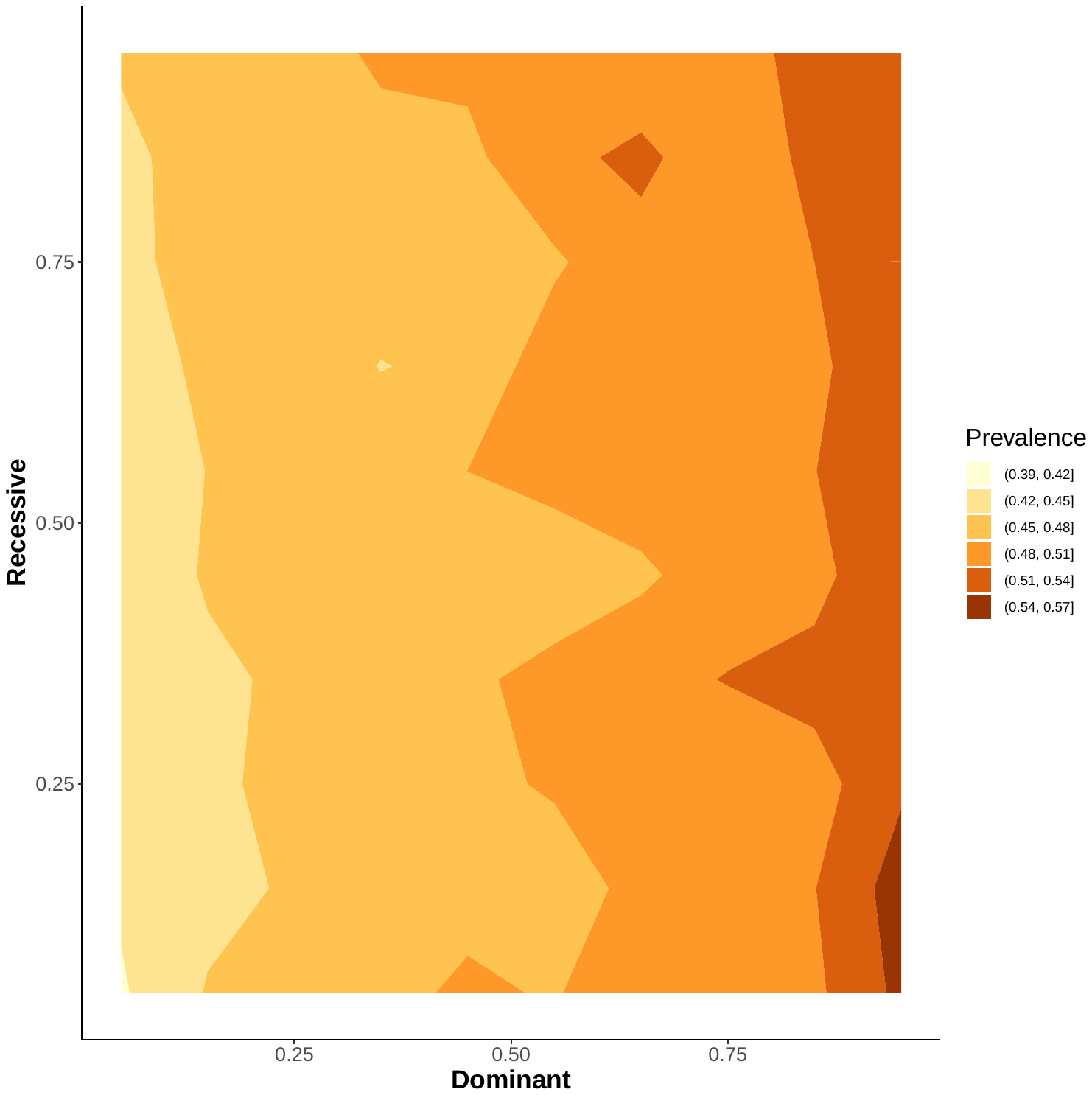 |
| c. Dominant and recessive PRS | |

Supplemental Figure 3. Multi-dimensional PRV and hypertension risk under p-value of 1.

| UKBB | eMERGE |
| --- | --- |
| 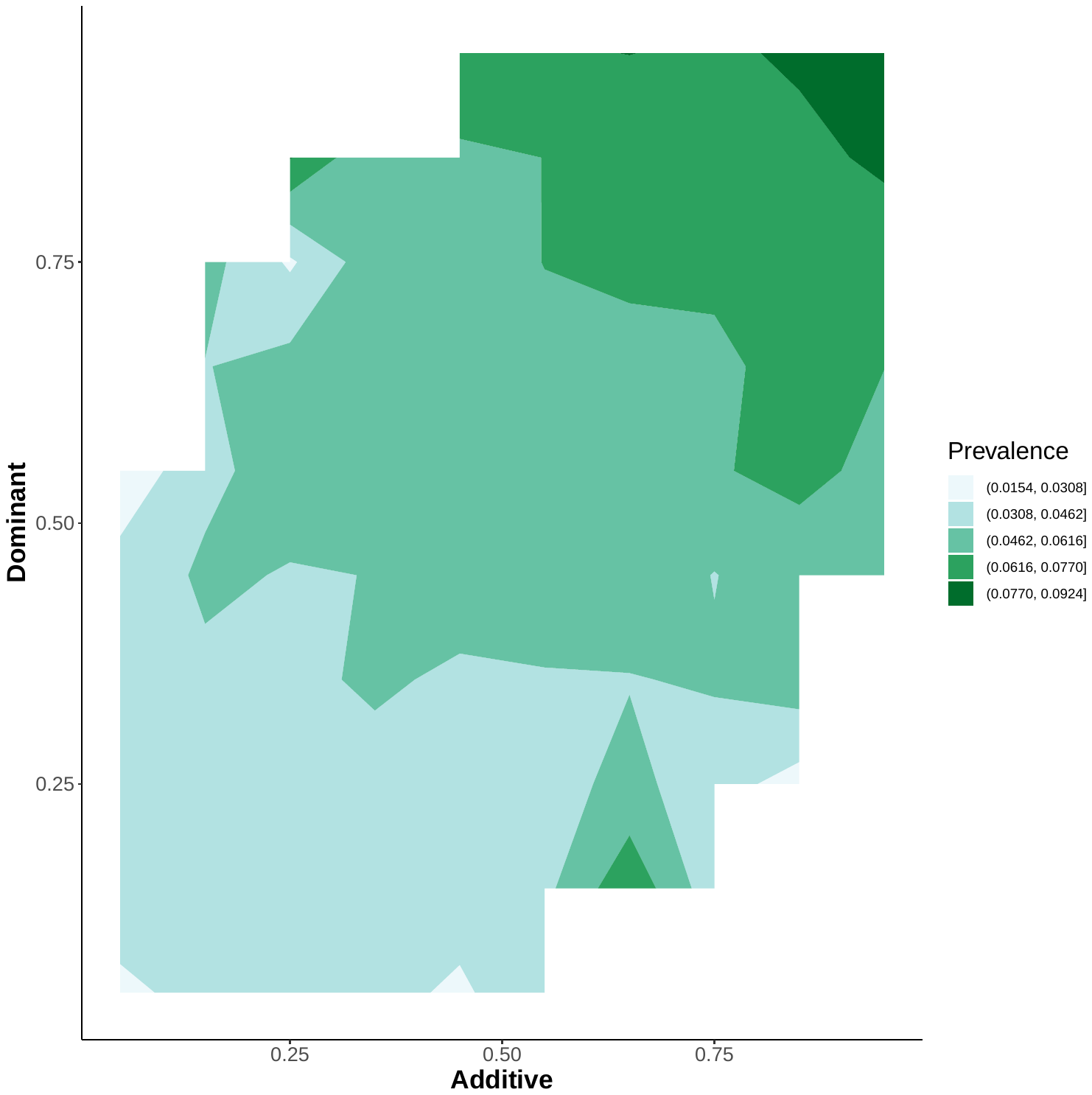 | 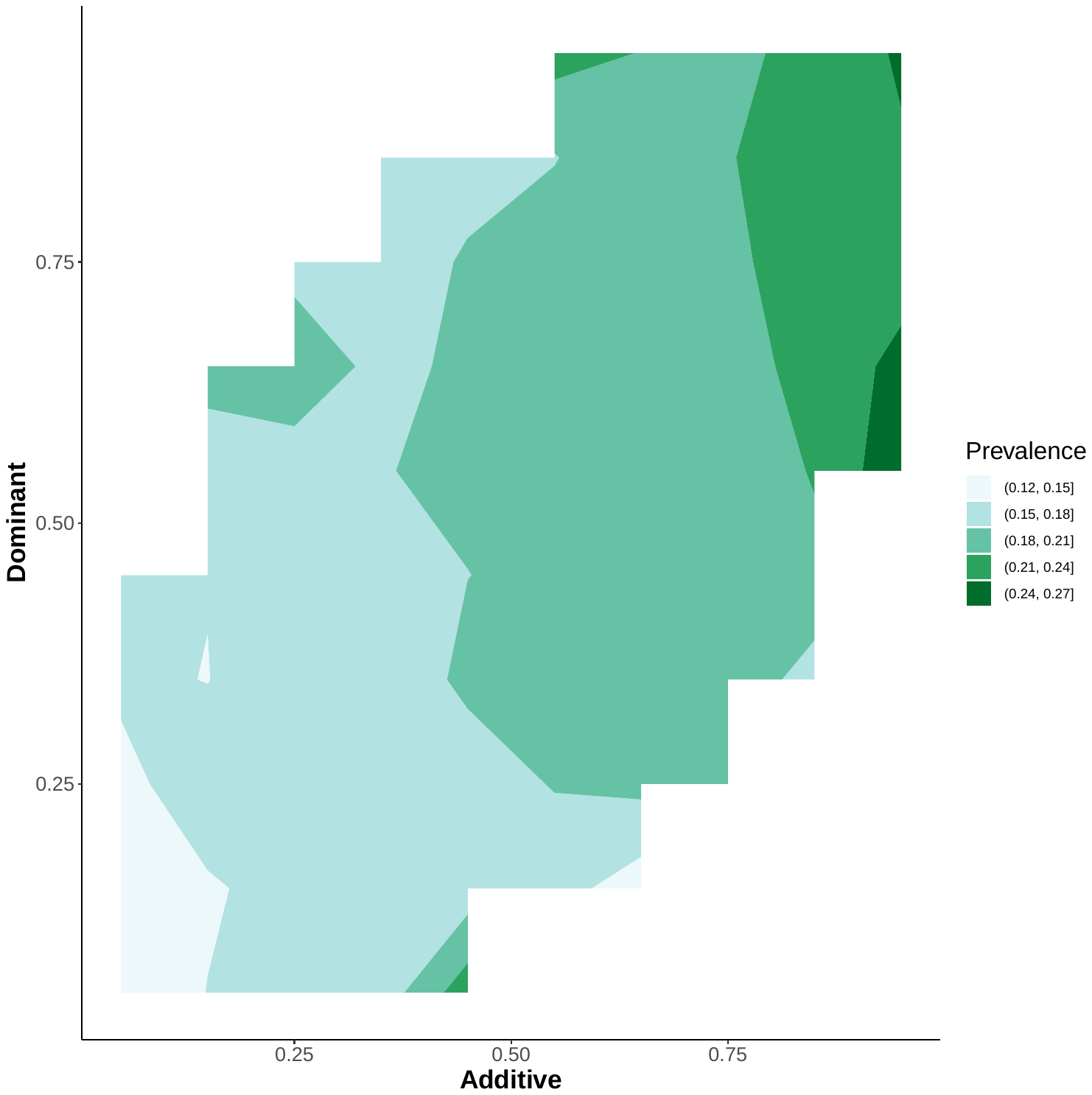 |
| a. Additive and dominant PRS | |
| 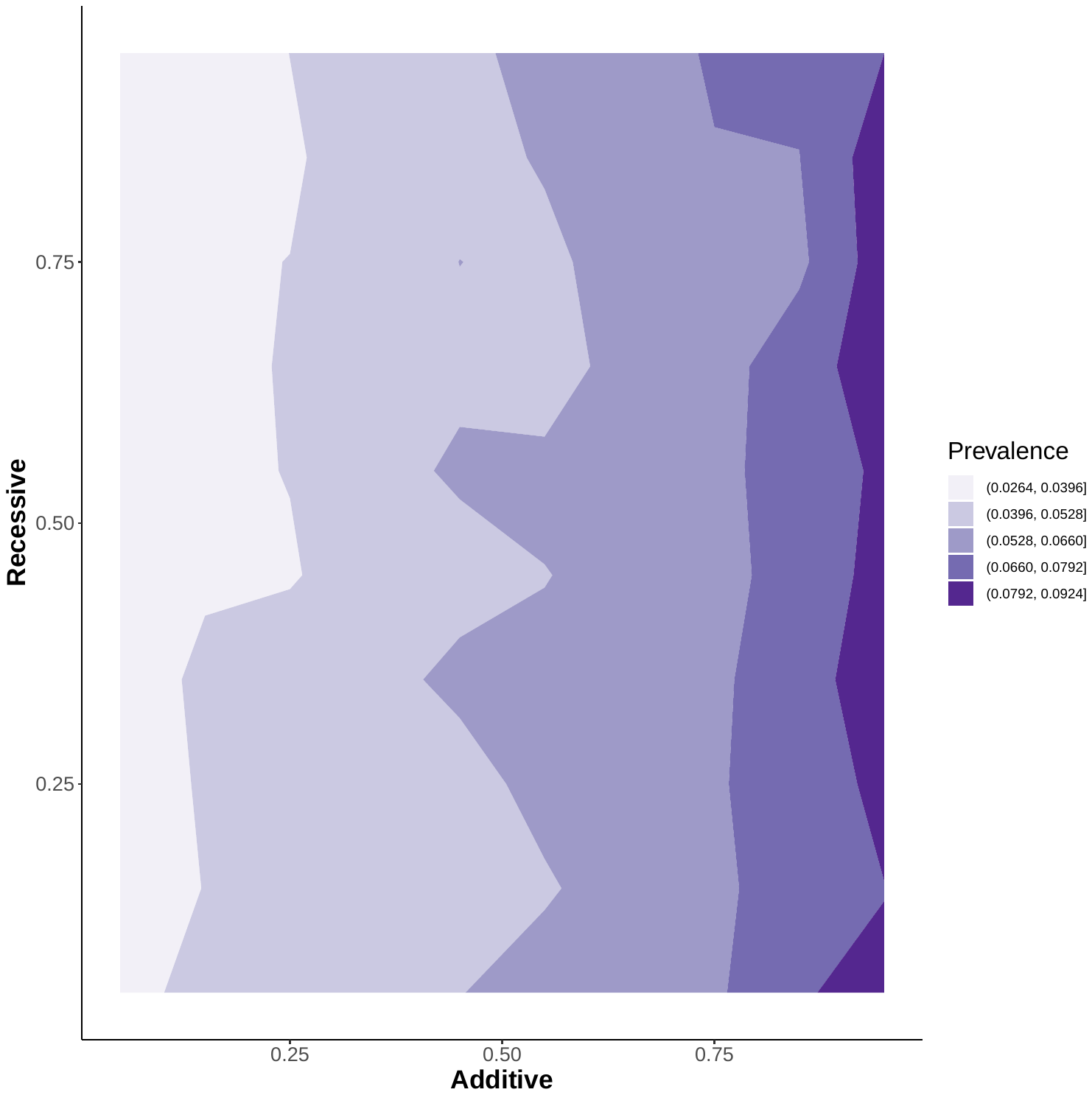 | 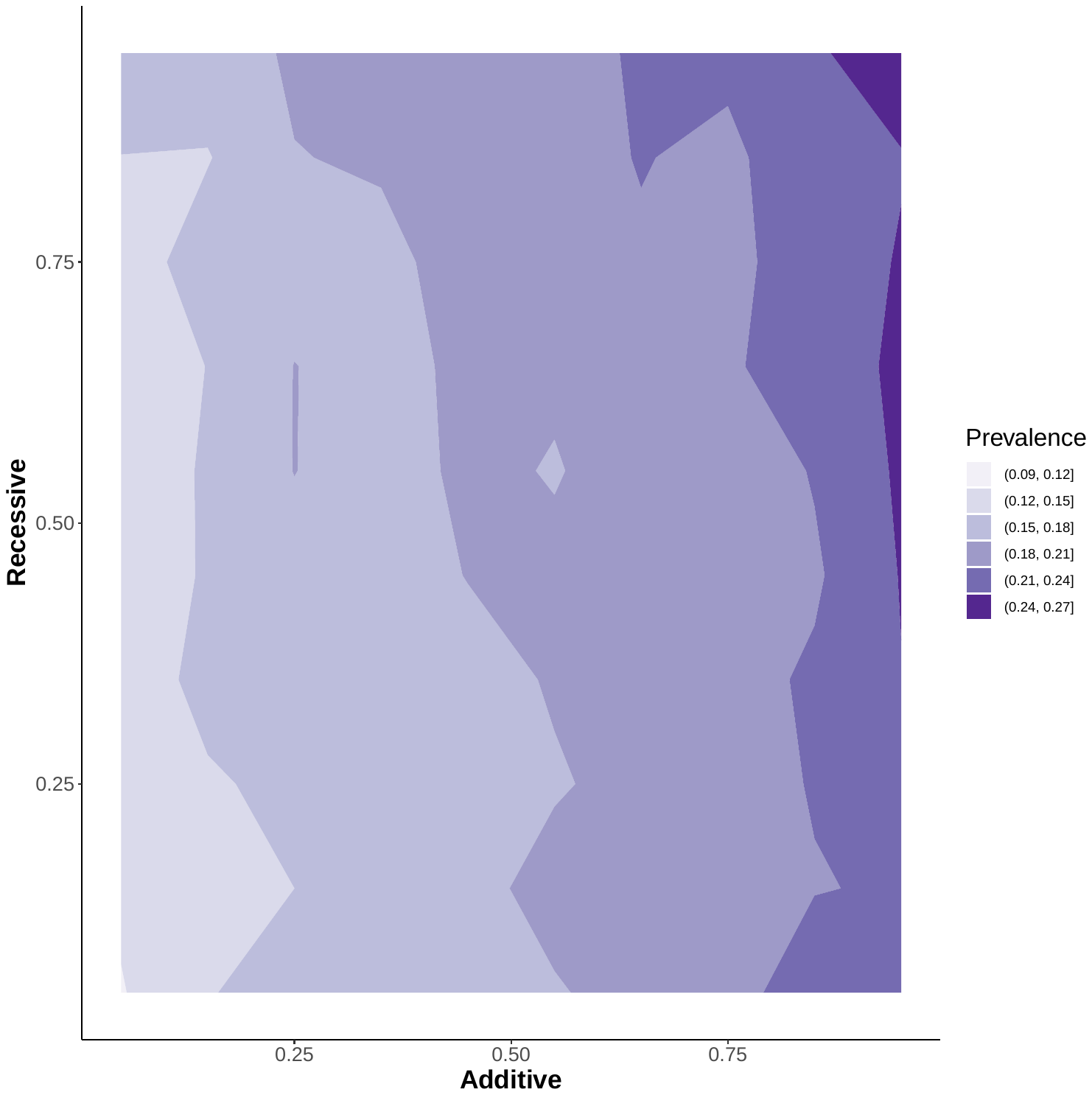 |
| b. Additive and recessive PRS | |
| 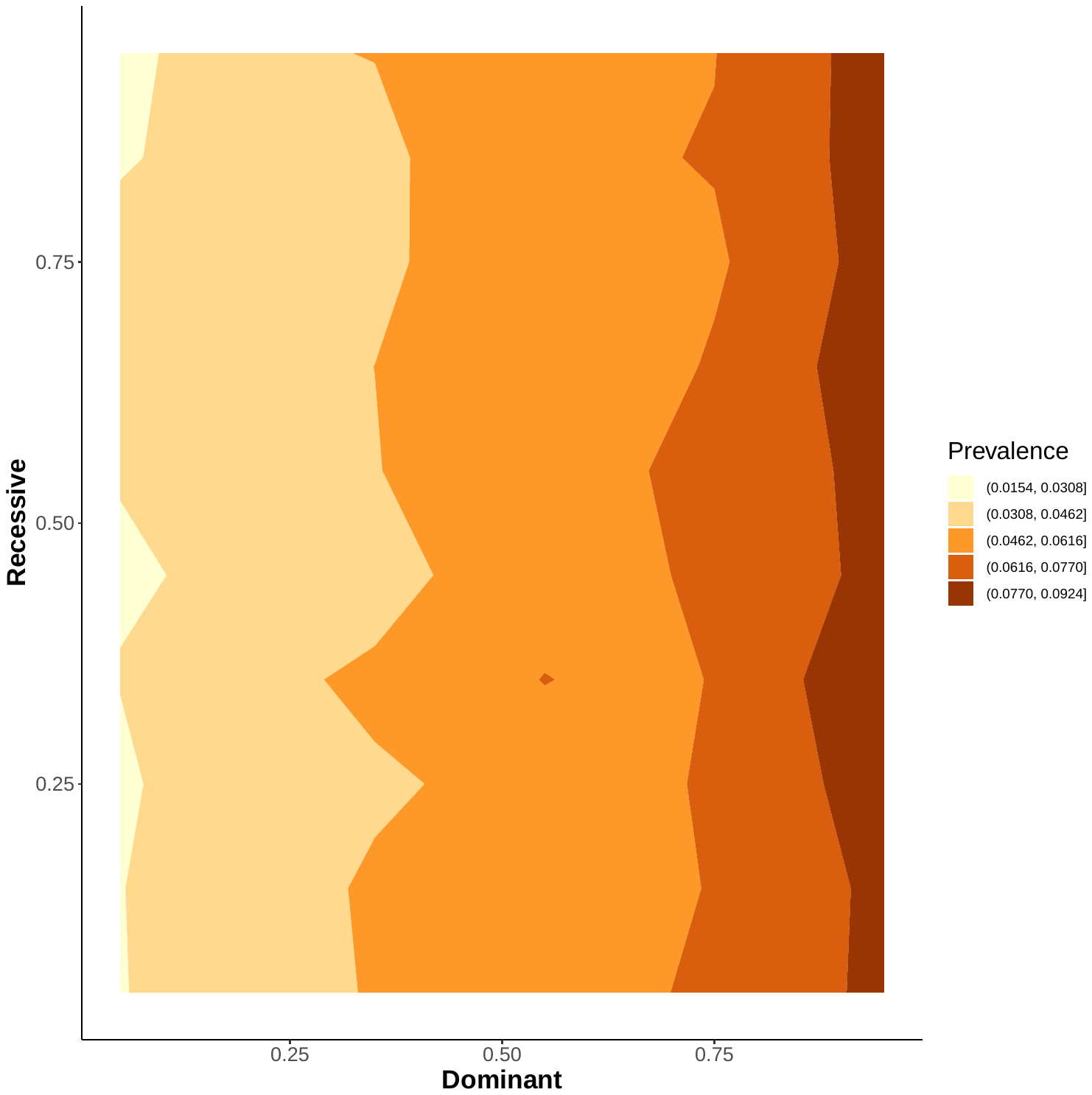 | 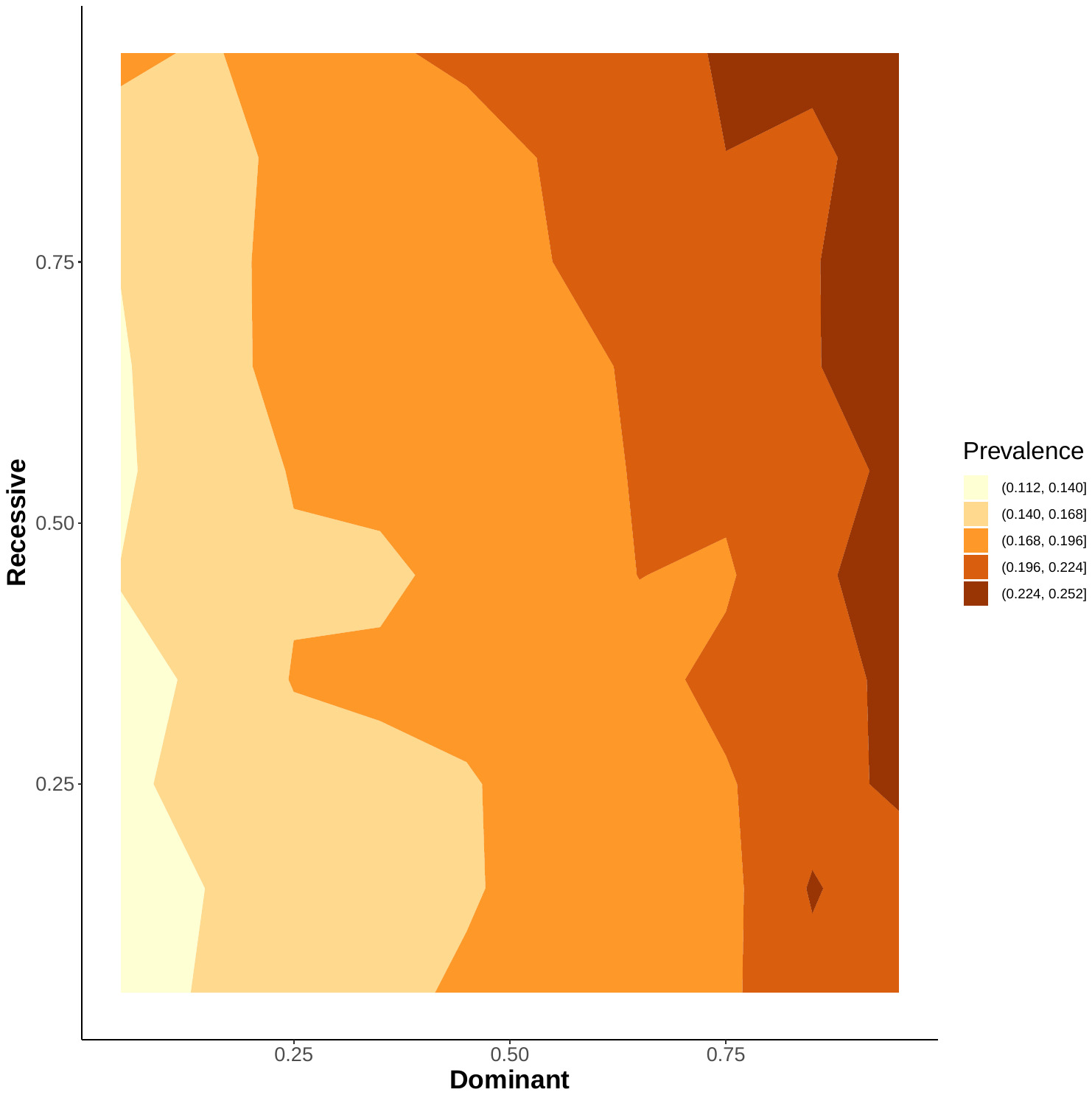 |
| c. Dominant and recessive PRS | |

Supplemental Figure 4. Multi-dimensional PRV and T2D risk under p-value of 0.05

| UKBB | eMERGE |
| --- | --- |
| 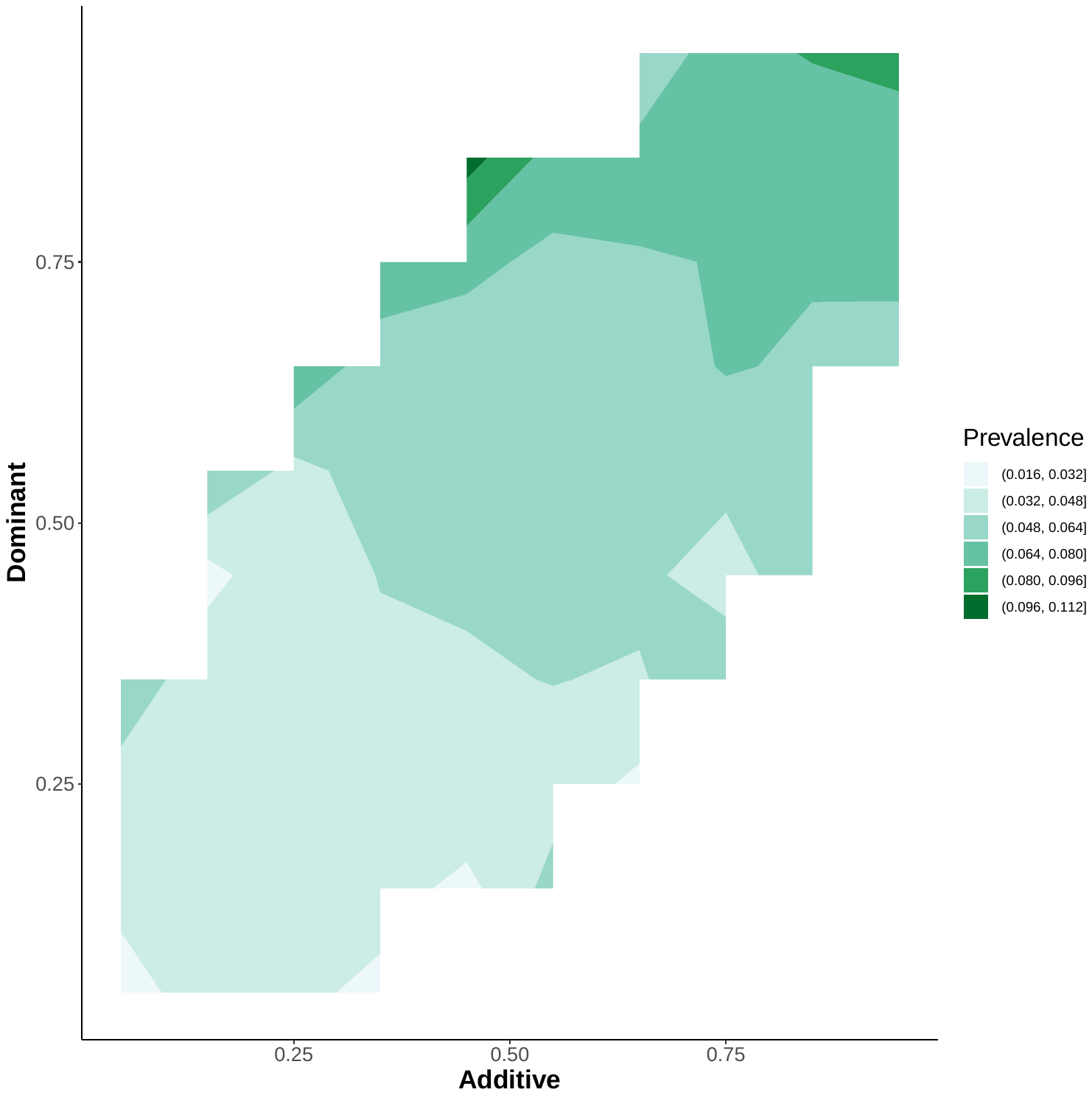 | 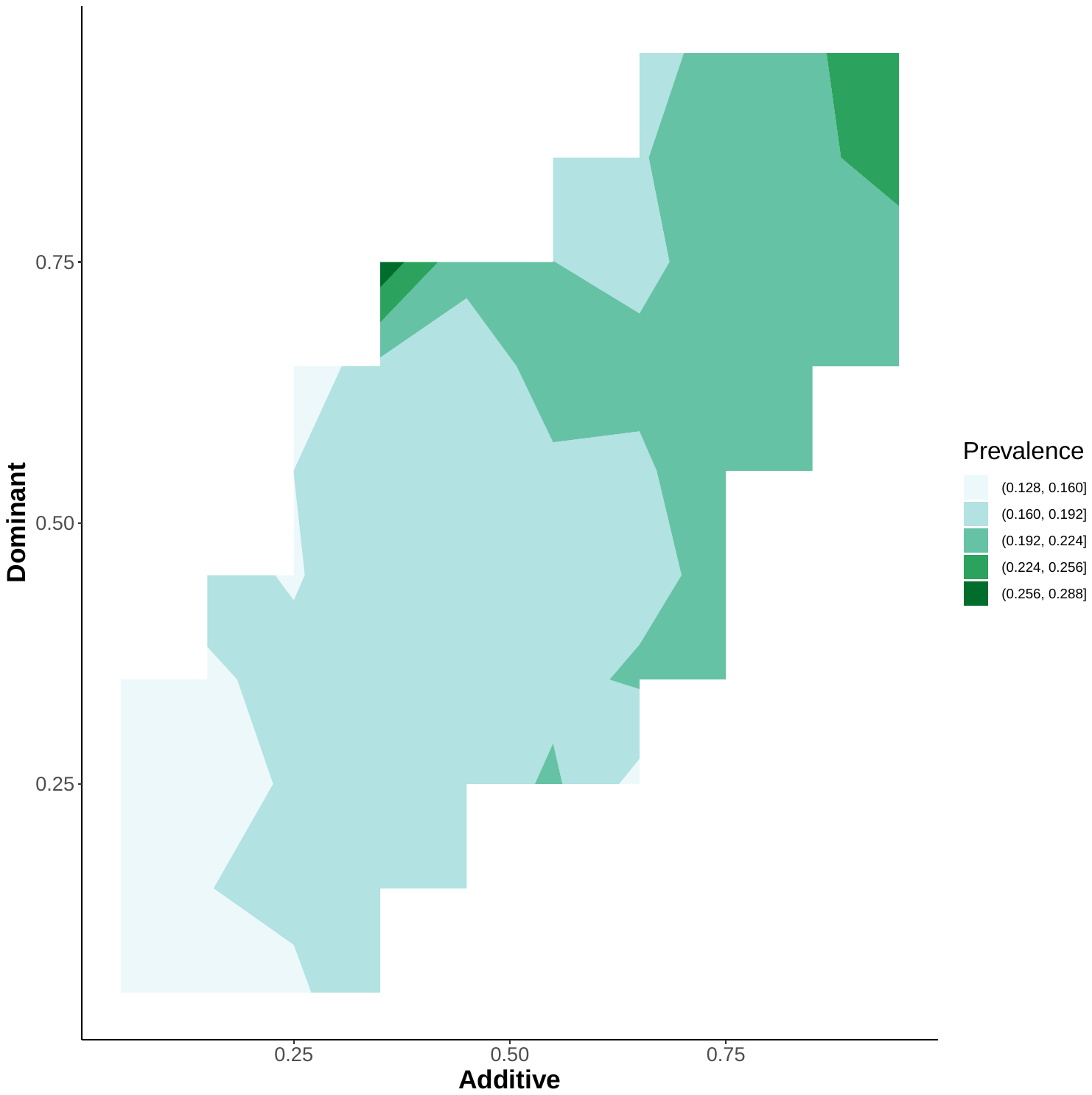 |
| a. Additive and dominant PRS | |
| 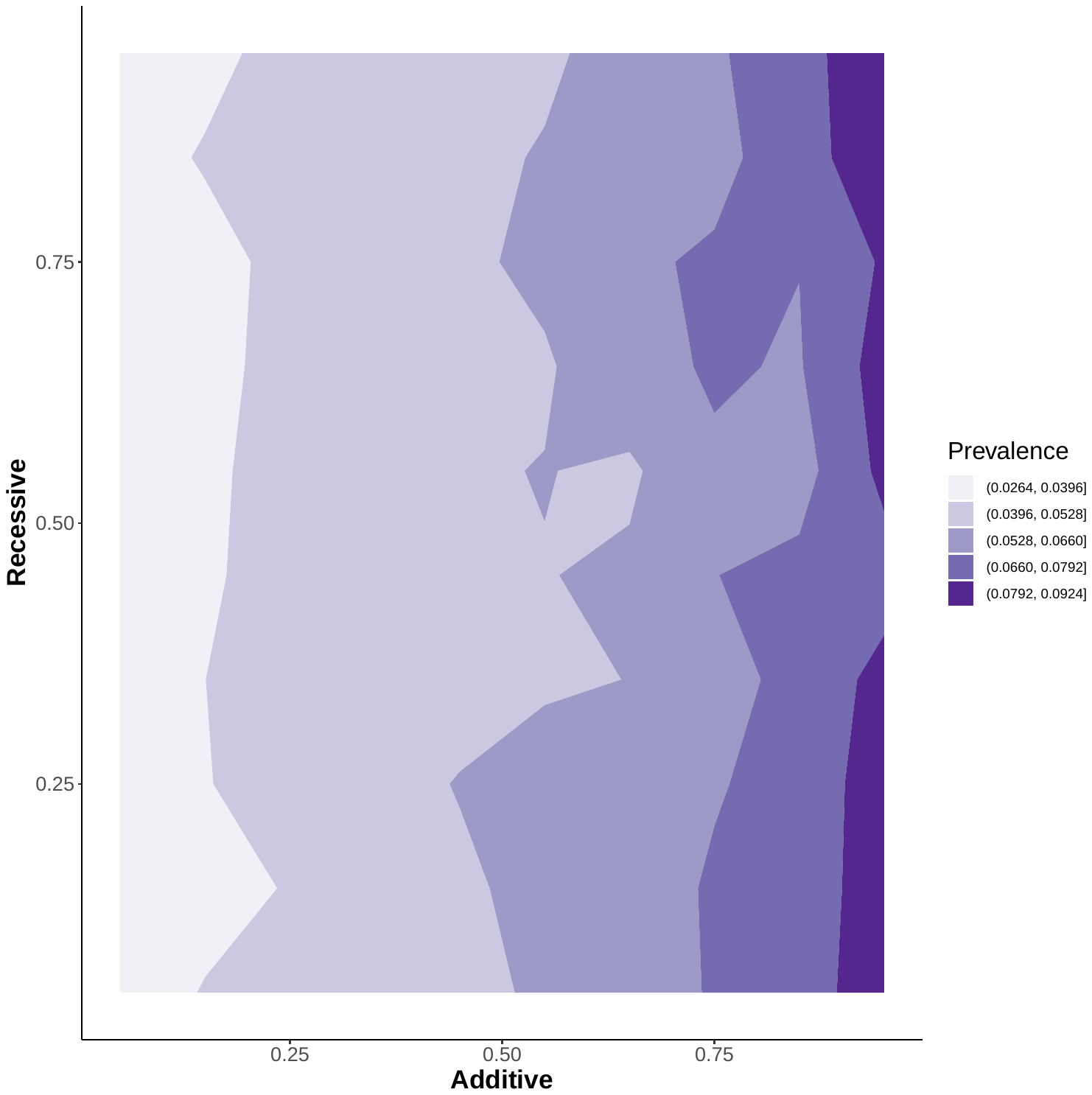 | 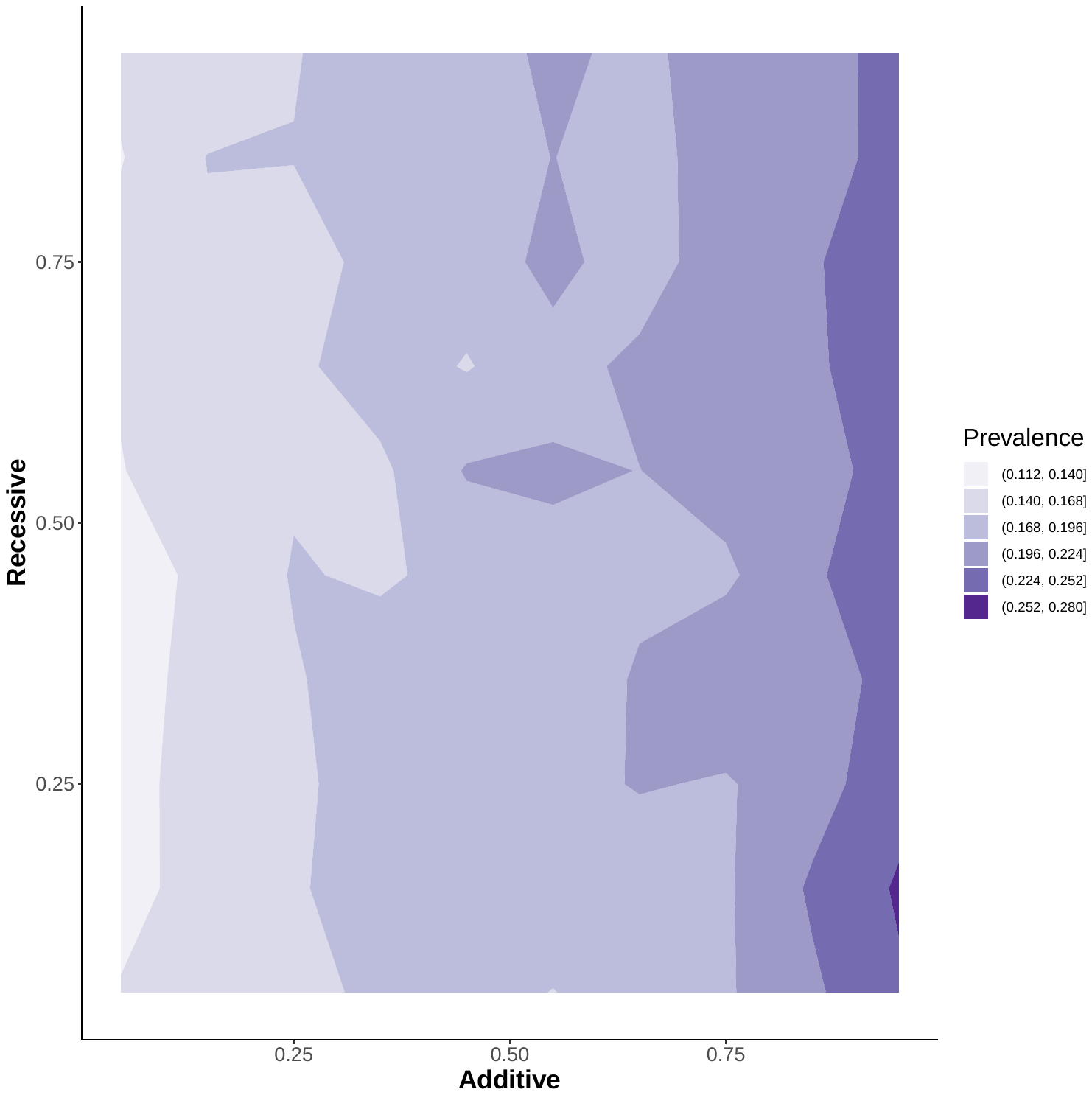 |
| b. Additive and recessive PRS | |
| 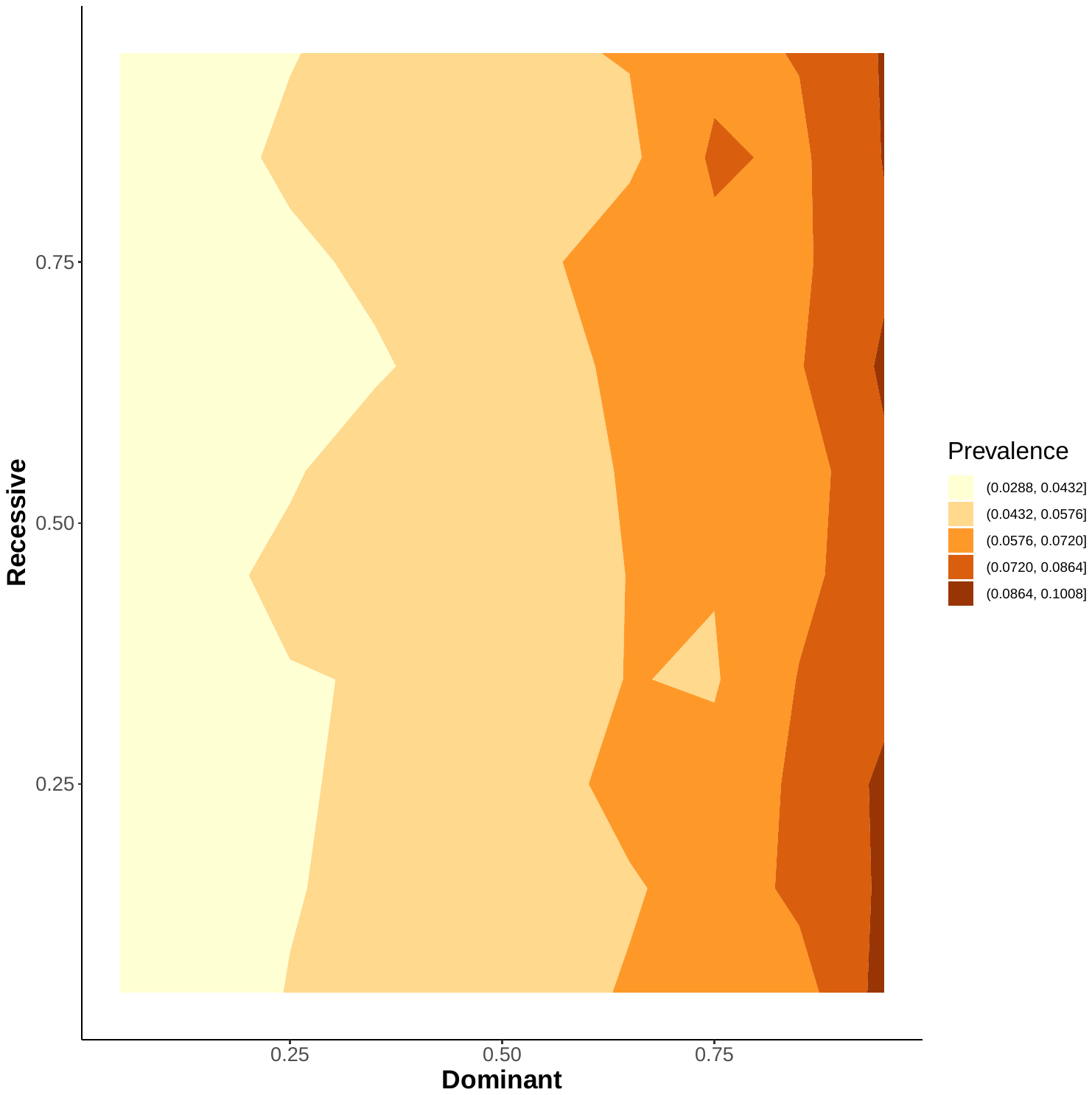 | 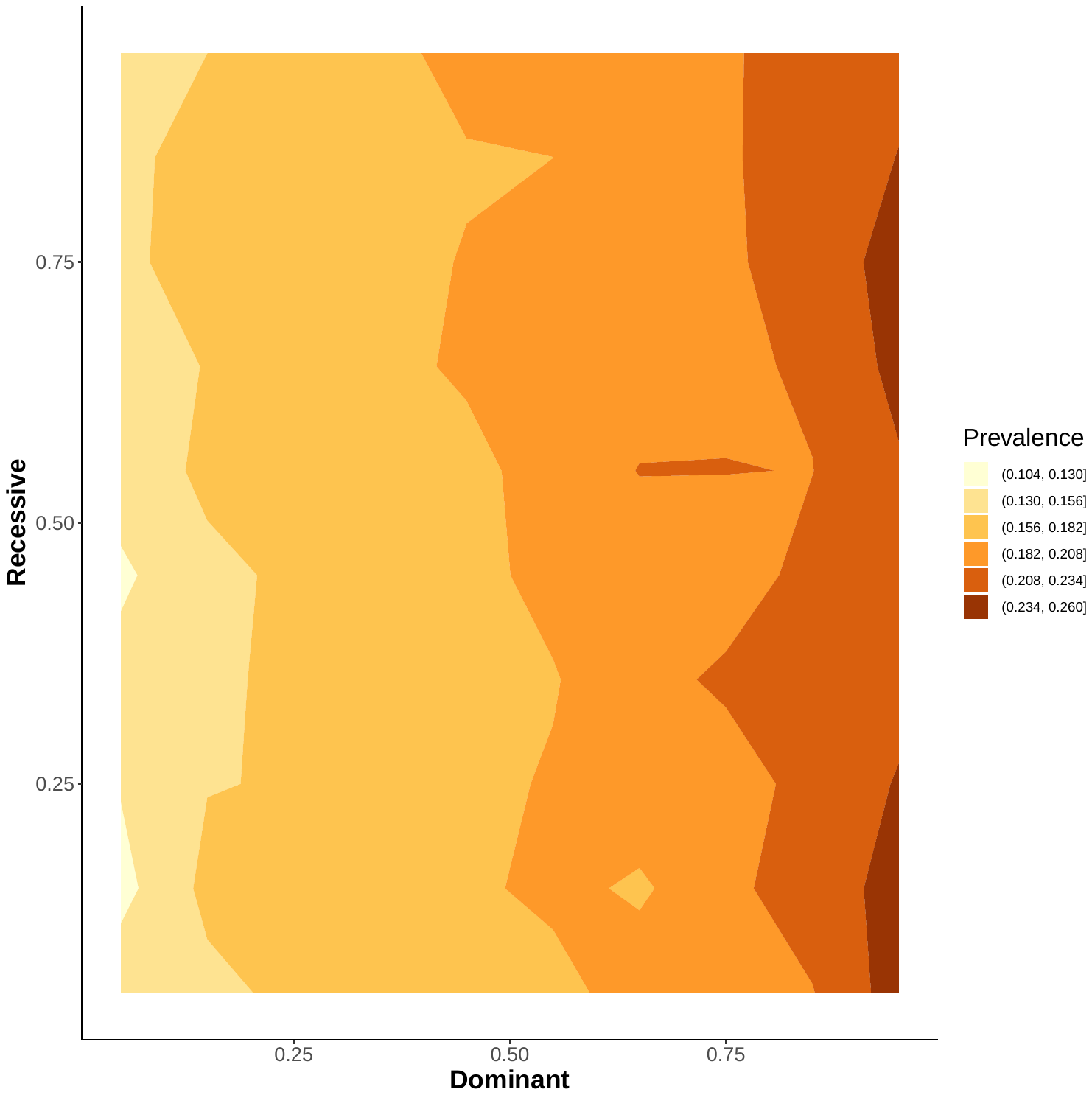 |
| c. Dominant and recessive PRS | |

Supplemental Figure 5. Multi-dimensional PRV and T2D risk under p-value of 1.
